# Effect of a needs-based model of care on the characteristics of healthcare services in England: the i-THRIVE National Implementation Programme

**DOI:** 10.1101/2024.05.07.24306984

**Authors:** R Sippy, L Efstathopoulou, E Simes, M Davis, S Howell, B Morris, O Owrid, N Stoll, P Fonagy, A Moore

## Abstract

**Aims:** Developing integrated mental health services that focus on the needs of children and young people is a key policy goal in England. The THRIVE Framework and its associated implementation programme, i-THRIVE, are now used in areas covering over 65% of England’s children. This study explores the experiences of staff involved with the i-THRIVE programme, assesses its effectiveness, and examines how local system working relationships influence the programme’s success.

**Methods:** The i-THRIVE programme was evaluated using a quasi-experimental study among twenty participating sites (ten implementation and ten comparison sites). Measurements included surveys of staff and leaders at each site and assessment of the “THRIVE-like” features of each site. Additional site-level characteristics were collected from health system reports. The effect of i-THRIVE was evaluated using a four-group propensity-score weighted difference-in-differences model; the moderating effect of local system working relationships was evaluated with a difference-in-difference-in-differences model.

**Results:** Staff at implementation sites were more likely to report using THRIVE in their own practice and exhibited better knowledge of THRIVE principles than comparison sites. The mean improvement among i-THRIVE sites was 16.7, and 8.8 among comparison sites. The results show that strong working relationships in the local system significantly enhances the effectiveness of the i-THRIVE programme. Sites with highly effective working relationships showed a notable improvement in “THRIVE-like” features, with an average increase of 16.41 points (95% confidence interval: 1.69–31.13, p-value: 0.031) compared to control sites. In contrast, sites with ineffective working relationships did not benefit from the i-THRIVE programme (−2.76, 95% confidence interval: −18.25–12.73, p-value: 0.708). This influence of working relationship effectiveness was consistent across various levels of THRIVE features.

**Conclusions:** The findings underscore the importance of working relationship effectiveness in the successful adoption and implementation of health policies like i-THRIVE.

## Background

Mental healthcare has numerous well-supported models. Globally, systems are challenged to provide appropriate services efficiently, necessitating reform of current care models (Hodgins et al., 2024). However, the continued presence of inadequate care is mainly due to difficulties in applying these changes (Shortell et al., 1993; Grol and Grimshaw, 2003), as systemic transformation is a complex process (Best et al., 2012). The transformative “Future in Mind” report (Department of Health, 2015) suggested deviating from a generic, tiered service model to a more responsive model for the specific mental health needs of local young populations. It called for service providers and funders to overhaul children and young people’s mental health (CYPMH) services into comprehensive systems, offering a range of services from prevention to risk management. The “Future in Mind” report focuses on reforming a flawed system of service provision, characterised by disunity, inefficiency, and limited access to services. Since 2016, Child and Adolescent Mental Health Services (CAMHS) within England’s National Health Service (NHS) have seen significant changes, advancing more comprehensive approaches to care (Rocks et al., 2020).

The THRIVE Framework provides a set of principles to guide system reformation, summarising the needs of CYP into five groups: Getting Advice, Getting Risk Support, Getting Help, Getting More Help, and Thriving (Wolpert et al.). Support is provided to CYP in these groups using a set of guiding principles. The THRIVE principles of care encompass characteristics of support at three levels: macro, meso, and micro. At the macro level, characteristics include interagency function and cooperation. This means a CAMHS system following the “THRIVE-like” approach would involve supporting bodies such as educational and social services in its policy-making and service delivery (Moore et al., 2023). At the meso level, they include a needs-based perspective focusing on CYP and their support services. A site adhering to meso-level THRIVE principles would be expected to have a network of community providers (Moore et al., 2023). At the micro level, these characteristics involve the interactions between CYP, their families, and healthcare professionals; we would expect to see shared decision-making, with everyone understanding the needs of the child and interventions being used (Moore et al., 2023).

The National i-THRIVE Programme (NIP) assists CAMHS sites in adopting the THRIVE principles, which prioritise patient needs and cohesive service provision through collaborative care networks (Moore et al., 2023). Over 65% of children and young people (CYP) live in areas where i-THRIVE has been adopted (i-THRIVE Team). i-THRIVE was created following implementation science guidelines to facilitate the adoption of THRIVE principles by CAMHS (Moore et al., 2016). The NIP is explained in a published study protocol (Moore et al., 2023). Briefly, i-THRIVE translates the complex aspects of a ‘THRIVE-like’ system into practical structures and tools for CAMHS to use in their transformation. The NIP guides CAMHS staff and leaders in developing local models based on THRIVE principles and creating detailed plans for implementation over four phases, using six components (Moore et al., 2023). The implementation strategies in the NIP are drawn from the Quality Implementation Framework (Meyers et al., 2012) and the Normalisation Processing Theory (May, 2006). The embodiment of THRIVE principles depends not only on efforts by the CAMHS site but also on broader community involvement (Wolpert et al.), requiring effective working relationships among local systems. According to the implementation strategy clusters created by the Expert Recommendations for Implementing Change study (Waltz et al., 2015), the implementation strategies used in the NIP include use of evaluative and iterative strategies, providing interactive assistance, adaptation and tailoring to context, development of stakeholder interrelationships, training and education of stakeholders, engagement of consumers, utilisation of financial strategies, and changes to infrastructure.

Evaluation of implementation strategies is needed to understand the impacts of the implementation and determine if it should be adopted for use in real-world scenarios. The evaluation could include several outcome metrics, such as acceptability, feasibility, fidelity or cost (Smith and Hasan, 2020). Fidelity is a particularly important metric, as it helps to determine if observed effects can be attributed to the intervention of interest; poor implementation may explain cases with no observed effects (Sanders et al., 2022). In this work, we evaluate the impact of the NIP implementation model on fidelity to the THRIVE principles of care; a healthcare service unit that embodies the THRIVE principles of care would exhibit high fidelity to THRIVE at macro, meso, and micro levels.

Although in use since 2016, the NIP as an implementation model has not been assessed. We focus on the average effect of NIP on the THRIVE fidelity of implementation sites. For this estimate, we need to know the outcome at each site if NIP had not been implemented (the potential outcome or unobserved counterfactual) (Stuart, 2010). The difference in differences (DiD) approach compares changes in outcomes between implementation and control sites before and after the intervention (Stuart et al., 2014). A key assumption for DiD is that the average outcomes for both groups would have similar trends over time (Xu, 2017), which may not always be plausible. Participation in NIP is voluntary, meaning sites that choose to participate may differ from those that do not. The four-group propensity score-weighted DiD method overcomes these issues by adjusting the implementation and control groups while considering time factors. This adjustment ensures comparability among the groups and reduces bias in estimating the desired effect. It’s particularly useful when group composition changes during the study, such as when health practitioners move between Clinical Commissioning Groups (CCGs).

In this study, we evaluate the alignment of frontline staff with THRIVE principles, and assess the effect of the NIP on THRIVE fidelity. We hypothesize that i-THRIVE had a positive effect on THRIVE fidelity. Additionally, we examine how the effectiveness of local system working relationships moderates the impact of the NIP on THRIVE fidelity.

## Methods

### Study Setting and Design

The study protocol is detailed elsewhere (Moore et al., 2023). In brief, we selected twenty CAMHS sites in England: ten of these sites have been using i-THRIVE since 2016 (NIP/implementation sites), and ten others were using different approaches for transformation (comparison sites). Details on the implementation strategies and delivery of NIP are in Supplemental Material S1. This study reporting conforms to the Template for Intervention Description and Replication (TIDieR) statement (Hoffmann et al., 2014); the TIDieR checklist is provided in Supplemental Material S2.

The NIP sites were Bexley, Cambridgeshire and Peterborough, Camden, Hertfordshire, Luton, Manchester, Stockport, Tower Hamlets, Waltham Forest, and Warrington. The comparison sites included Bradford, Ipswich and East Suffolk, Lewisham, Norfolk, Northampton, Portsmouth, Southampton, Stoke-on-Trent, Sunderland, and South Worcestershire. Sites will be pseudonymised as Site A—T. Additional site details are in Supplemental Material S3. Implementation of THRIVE started in April 2016; data from April 2015 to March 2016 were used as the pre-implementation period. Data from April 2018 to March 2019 were used as the post-implementation period. To examine staff alignment with and understanding of THRIVE principles, a survey was administered. To assess site transformation or implementation, site leaders were also surveyed. To assess whether THRIVE principles were embraced by sites, fidelity scores were assigned. To understand whether the NIP impacted site adoption of THRIVE principles, fidelity scores from pre- and post-implementation were compared between implementation and comparison sites, while adjusting for site-level characteristics (auxiliary data).

### Measurements and Data

#### Surveys

Two surveys were designed based on the RE-AIM Adoption Framework (Glasgow et al., 2019), a guide for evaluation of programmes. Both surveys were primarily quantitative in nature, but several questions requested open-ended responses. Copies of the surveys are in Supplemental Materials S4—S6. The staff survey assessed the staff awareness of THRIVE and use of THRIVE principles. Site leaders distributed the survey to professionals involved in providing and commissioning CYPMH services at both i-THRIVE and comparison sites. The survey was administered in August 2019, October 2019, and January 2020.

A survey for transformation leads was also conducted among i-THRIVE and comparison sites. This survey, completed by programme managers, gathered information about site transformation activities and the support they received. It was distributed from January to May 2020. Additional details on the surveys are in Supplemental Material S3.

#### Principal Outcome: Fidelity

The primary focus is the degree to which sites follow the THRIVE principles. The THRIVE principles of care encompass macro-, meso-, and micro-level features. We assessed these features in participating sites using the i-THRIVE Assessment Tool (Moore et al., 2023), wherein a score is assigned to each of 75 items describing THRIVE principles (1: low, 4: high). Purposive sampling was used to recruit three staff members at macro (senior leadership), meso (service management), and micro (frontline staff) levels for in-depth interviews. Interview transcripts were scored by evaluators: sites received an overall fidelity (300 possible) and level-specific scores (macro: 84 possible, meso: 104 possible, micro: 112 possible). Higher scores indicate better adherence to THRIVE principles. Fidelity to i-THRIVE was measured at each site before and after the intervention by at least two independent evaluators, with scores averaged for each site and period. When there was not enough information to assign a score, we estimated the missing scores using the mean difference between pre- and post-intervention scores for the site, based on the score for the available period. Additional details on fidelity ratings are in Supplemental Materials S3.

#### Auxiliary Data

To calculate propensity score weights, it is important to identify site characteristics that might cause selection bias or have a confounding effect (Stuart et al., 2014). These characteristics were measured in 2016 and 2019, unless stated otherwise. They included population density (total population per square kilometre) for each CCG (Office for National Statistics, 2016; Office for National Statistics, 2017), annual funding support (£100,000 increments) per CCG (NHS England, 2017), Indices of Multiple Deprivation (IMD) ranks (Department for Communities and Local Government, 2015; Department for Communities and Local Government, 2019), the initial number of CCGs per site (NHS England, 2019), and compliance in 2017 with the CCG Improvement and Assessment Framework Mental Health Transformation Milestones (NHS England, 2017). For sites comprising multiple CCGs, we summed the annual funding and averaged the population density, effectiveness of working relationships in the local system, and transformation compliance across CCGs. Site-level IMD ranks were calculated using the method recommended by the Office of National Statistics (Noble et al., 2019).

### Statistical Analysis

We analysed the staff survey results using the chi-squared test and Cramer’s V as a measure of association between i-THRIVE/comparison sites and staff responses (“yes”/”no”). We used a binomial distributed generalised linear model with a log link for site-specific results, excluding “I don’t know” responses. To determine if any site had an unusual probability of a “yes” response, we compared the “yes” probability for each site to the average among comparison sites.

The responses from the survey of the implementation leads are in Supplemental Material S1. Their experience of the delivery of i-THRIVE was summarised as proportions. To assess inter-rater reliability for the fidelity scores, we used Krippendorf’s alpha (see Supplemental Material S3 for more details).

Voluntary participation in health policy implementations like the NIP can lead to selection bias, as the sites that choose to participate may differ from those that do not. To correct for this, we applied propensity score weighting (Rosenbaum and Rubin, 1985) to equalise the distribution of characteristics between the implementation and comparison sites. We used a four-group weighting method (pre-implementation i-THRIVE, post-implementation i-THRIVE, pre-implementation comparison, and post-implementation comparison) to align characteristics across all groups with those of the pre-implementation i-THRIVE sites (Stuart et al., 2014). These propensity scores were calculated using a multinomial model with five site characteristics (population density, annual funding, IMD rank, the number of CCGs per site, and transformation compliance). Characteristic balance was checked using the standardised difference in means (Stuart, 2010). The impact of NIP on fidelity was estimated using maximum-likelihood repeated measures linear regression with an auto-regressive correlation structure, weighted with the calculated propensity scores. To account for remaining characteristic imbalances, we included population density, IMD, and transformation compliance in the final model. The results from this model represent the four-group weighted DiD effect estimate.

To assess the reliability of our fidelity results, we conducted a series of sensitivity analyses. We employed different methods to estimate the effect of the NIP, including the standard (unweighted) DiD and alternative model specifications. We examined the impact of non-compliant control sites by repeating the analysis, excluding these sites. More details can be found in Supplemental Material S7.

To examine variations in the effect of i-THRIVE, we investigated a possible effect moderator, specifically the quality of local system working relationships (NHS England, 2021) on i-THRIVE implementation. Further details are available in Supplemental Material S9.

All data visualisation, cleaning, and propensity score modelling was performed in R version 4.2.3 (The R Foundation for Statistical Computing, Vienna, Austria) and RStudio version 2023.03.0 (RStudio, Inc., Boston, MA, USA), with R packages cowplot, extrafont, ggpattern, ggspatial, gridExtra, haven, irr, nnet, RColorBrewer, readxl, reshape2, sf, tableone, and tidyverse (Venables and Ripley, 2002; Wickham, 2007; Neuwirth, 2014; Auguie, 2017; Wickham and Miller, 2017; Pebesma, 2018; Gamer et al., 2019; Wickham et al., 2019; Wilke, 2020; FC et al., 2022; Yoshida and Bartel, 2022; Chang, 2023; Dunnington, 2023; Pebesma and Bivand, 2023; Wickham and Bryan, 2023). The simple DiD, four-group weighted DiD, and effect modification analyses were performed in SAS® version 3.81 (SAS Institute Inc., Cary, NC, USA) in SAS® Studio.

## Results

### Surveys

The staff survey had 689 responses across 19 sites (no responses from Luton). Detailed survey results are in Supplemental Material S8.

While the THRIVE Framework was widely known, more implementation respondents recognised it (83.9% vs. 70.5%, p<0.0001). A higher proportion of implementation respondents reported using THRIVE principles in their daily practice (58.5% vs. 49.0%, p=0.03). Implementation respondents were more likely to score perfectly on a test of the THRIVE Framework (34.1% vs. 22.9%, p=0.001).

The transformation leads survey included eight managers from seven implementation sites and eight managers from seven comparison sites. Notably, managers from four comparison sites (Sites E, J, K, and S) reported using THRIVE as their service transformation model. This prompted a more detailed examination of staff survey results concerning THRIVE implementation at comparison sites.

The site-level analysis of survey results can be found in Supplemental Material S8. Among those who provided a yes/no response at comparison sites, J and K reported a higher likelihood of implementing THRIVE. The odds of respondents reporting site implementation of THRIVE were 4.43 in J (95% CI: 2.32–8.47) compared to other comparison sites (76.5% vs. 59.5%), and 4.43 in K (95% CI: 1.33–14.80) compared to other comparison sites (76.5% vs. 59.5%). Among respondents at comparison sites, there was no difference in personal use of THRIVE principles compared to other comparison sites. Regarding knowledge of the THRIVE Framework, respondents from J had a higher probability of achieving a perfect score on the quiz: the odds of scoring perfectly were 3.73 (95% CI: 2.25–6.19), compared to other comparison sites (37.7% vs. 22.9%).

### THRIVE Fidelity

During the pre-implementation period, i-THRIVE sites had an average fidelity score of 149.0 (range: 132.0—180.2) and comparison sites had an average score of 133.4 (range: 113.0— 158.2). Following implementation, i-THRIVE sites had an average score of 166.6 (range: 145.5—195.0), while comparison sites had an average of 142.2 (range: 132.0 —175.0). The mean difference between pre- and post-implementation among i-THRIVE sites was 16.7; among comparison sites the mean difference was 8.8. Two sites had incomplete fidelity score information: the macro-level components for Site T during the post-implementation period and the meso-level components for Site F during the pre-implementation period (scores were assigned as outlined in the methods section). Detailed fidelity scores by level and site are illustrated in Figure 1, and a map showing the changes in scores by site throughout the study is presented in Figure 2.

**Figure 1:**
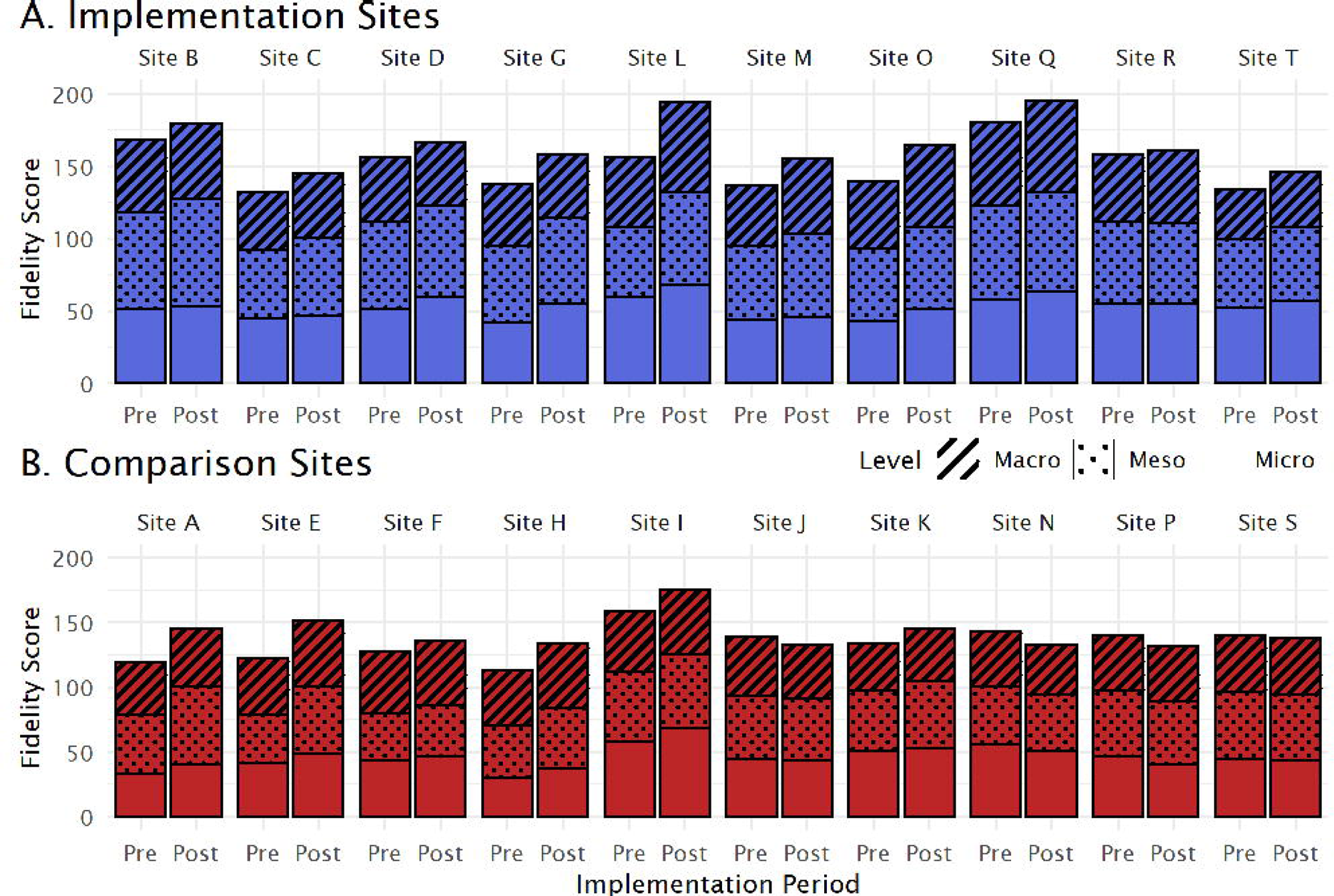
Fidelity Scores by Site. Total fidelity scores during the pre- and post-implementation periods are represented by bar height, with patterned overlay to indicate component levels (macro, meso, micro). Implementation sites are in panel A (blue) while comparison sites are in panel B (red).

**Figure 2:**
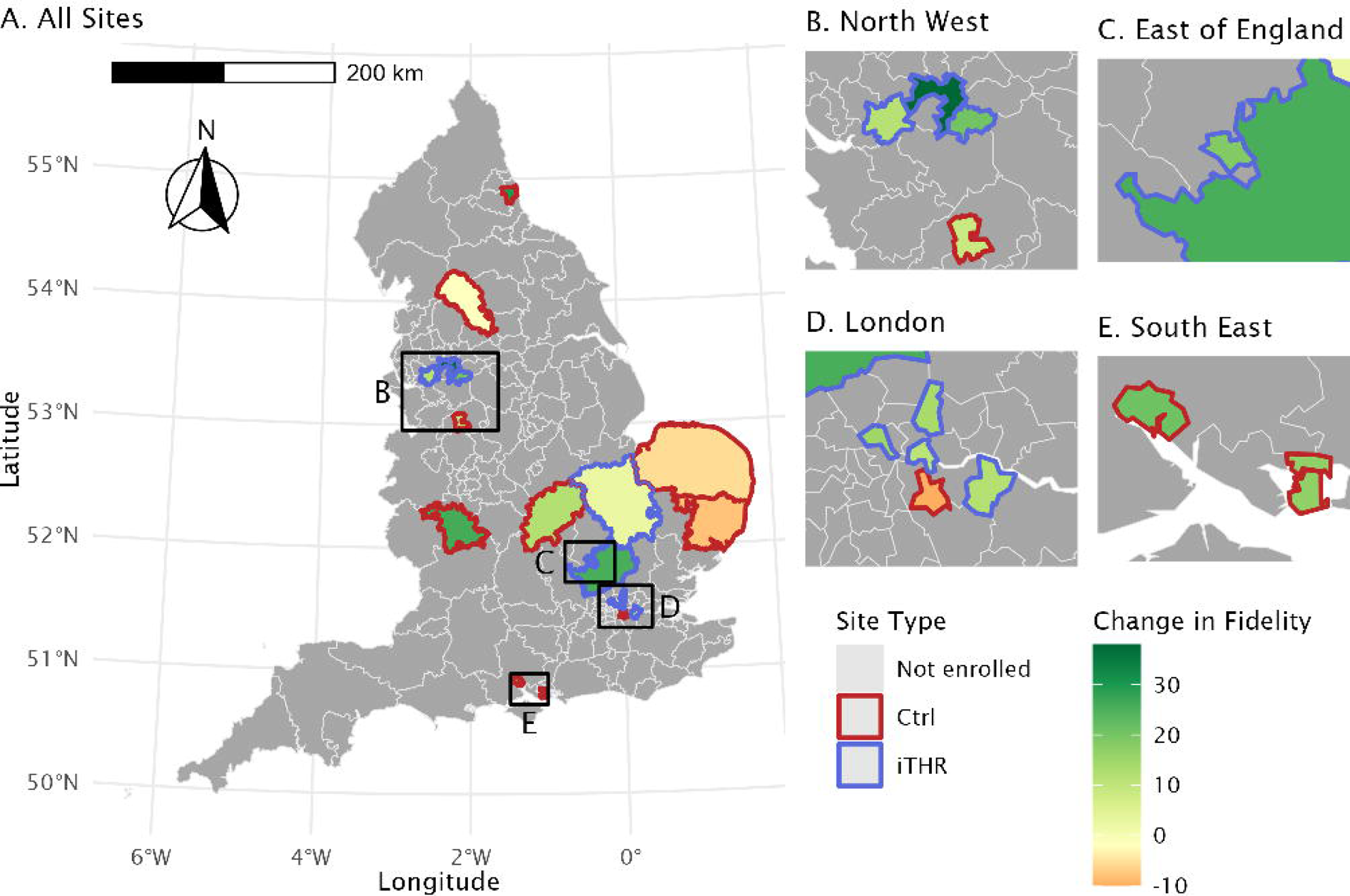
Change in Fidelity Scores Over Study Period. The difference in total fidelity scores during the pre- and post-implementation periods are represented by colour, with an increased score in green and a decreased score in red, on a background map of Clinical Commissioning Groups in panel A. Study sites have a bold outline (comparison sites in red, implementation sites in blue). Inset maps for North West/Midlands, London, and South East are in panels B–D, respectively.

Site characteristics, adjusted using four-group propensity-score weighting, are in Table S6, Supplemental Material S8. In our analysis of the weighted standardised differences, we identified some remaining imbalances, specifically in population density for the pre-implementation control group, IMD rank for both control groups, and transformation compliance for both control groups. These covariates were included in our effect estimate model, ensuring a more accurate assessment of NIP impact.

The NIP effect estimates are presented in Table 1. Our analysis reveals that the overall fidelity scores were moderately influenced by the NIP. Specifically, i-THRIVE sites showed an average improvement of 7.05 points (95% CI: −4.47–18.57). The most notable improvements were at the macro level, where i-THRIVE sites increased by an average of 2.92 points (95% CI: −1.09–6.92), followed by the meso level with an average increase of 2.76 points (95% CI: −1.98–7.51), and the micro level with an average increase of 1.39 points (95% CI: −3.94–6.72). None of these improvements were statistically significant. When comparing the four-group weighted DiD with the standard DiD analyses (Supplemental Material S7), we found comparable effects on overall and macro-level fidelity. There were shifts in the impacts on meso- and micro-level fidelity. This suggests that lower-level fidelity was more sensitive to the disparities between i-THRIVE and comparison groups, which were corrected through the four-group propensity-score weighting approach. Alternative modelling approaches produced results similar to our analysis. The exclusion of non-compliant comparison sites (i.e. Site J; see Supplemental Material S7) did not alter the results.

**Table 1:**
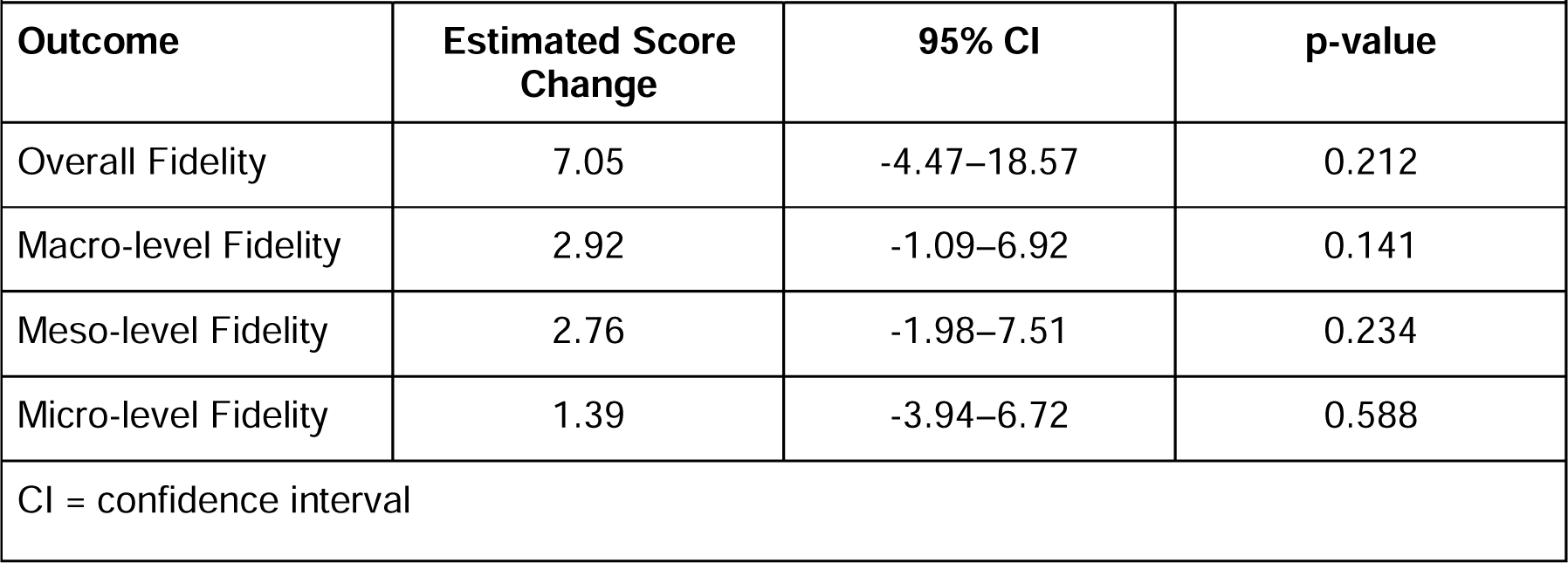
Estimates of Association Between the National i-THRIVE Programme and THRIVE Fidelity.

Among the study sites, five implementation sites and five comparison sites were found to have highly effective working relationships among their local systems (Supplemental Material S9). We observed a moderating effect of local systems working relationship effectiveness on the impact of the NIP for overall and macro-level fidelity. i-THRIVE was found to be more effective at sites with highly effective working relationships. The detailed results are in Table 2. i-THRIVE sites with highly effective working relationships showed increased fidelity scores compared to comparison sites with highly effective working relationships. The most significant impact was on overall fidelity scores (16.41, 95% CI: 1.69–31.13), followed by macro-level scores (6.95, 95% CI: 2.15–11.75). The moderating influence of highly-effective working relationships on meso-level and micro-level fidelity was modest (5.52, 95% CI: −0.66–11.71 and 3.95 points, 95% CI: −3.24–11.15, respectively). Notably, there was no discernible impact of the NIP on sites with ineffective working relationships across any fidelity level.

**Table 2:**
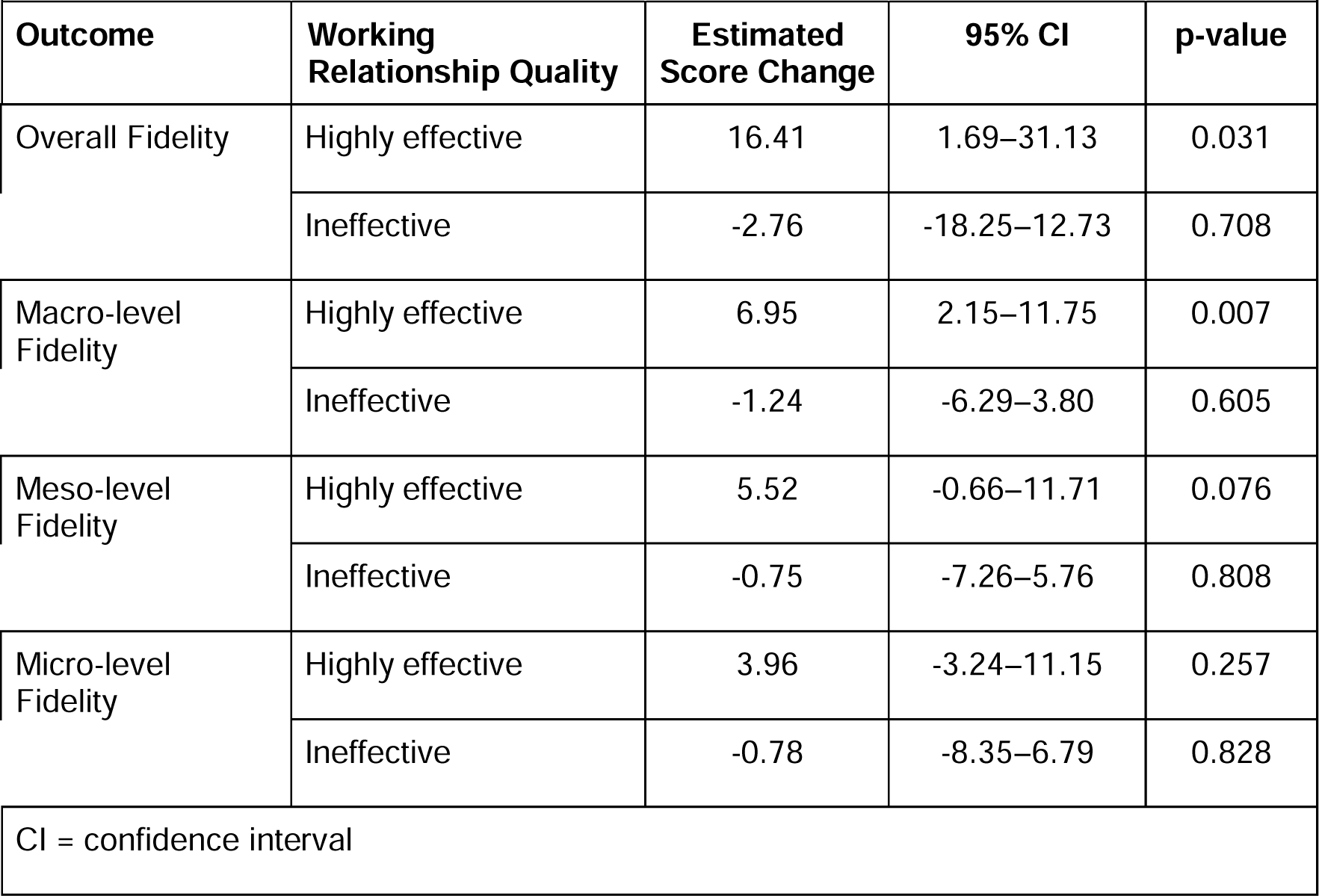
Effect Modification by Working Relationship Quality on the Impact of the National i-THRIVE Programme.

|  |  |  |  |  |
| --- | --- | --- | --- | --- |
| **Outcome** | **Working Relationship Quality** | **Estimated Score Change** | **95% CI** | **p-value** |
| Overall Fidelity | Highly effective | 16.41 | 1.69–31.13 | 0.031 |
|  | Ineffective | -2.76 | -18.25–12.73 | 0.708 |
| Macro-level Fidelity | Highly effective | 6.95 | 2.15–11.75 | 0.007 |
|  | Ineffective | -1.24 | -6.29–3.80 | 0.605 |
| Meso-level Fidelity | Highly effective | 5.52 | -0.66–11.71 | 0.076 |
|  | Ineffective | -0.75 | -7.26–5.76 | 0.808 |
| Micro-level Fidelity | Highly effective | 3.96 | -3.24–11.15 | 0.257 |
|  | Ineffective | -0.78 | -8.35–6.79 | 0.828 |
| CI = confidence interval |  |  |  |  |

## Discussion

A major obstacle in implementing any complex intervention is ensuring it becomes a routine practice across the entire organisation. This concept is often described as making it ‘the way we do things around here’ (Haines et al., 2004; Proctor et al., 2011). Both THRIVE and i-THRIVE are frequently mentioned in child mental health policy documents, by NHS England’s regional transformation boards, and in media related to child mental health. It is encouraging for policymakers to note that over 70% of staff at comparison sites were aware of THRIVE, and nearly 23% exhibited perfect knowledge of THRIVE principles. At the site level, respondents from most i-THRIVE sites had an increased odds of reporting site implementation of THRIVE, compared to the average among comparison sites, though two comparison sites (Sites J and K) also had an increased odds of reporting site implementation of THRIVE. When asked about personal use of THRIVE principles, respondents from many i-THRIVE sites had an increased odds of reporting personal use, compared to the average among comparison sites, and with Site J also exhibiting an increased odds of reporting personal use. This motivated a sensitivity analysis in which Site J was excluded from the fidelity analysis; the results were not affected. Overall, staff at i-THRIVE sites were significantly more likely to be familiar with, understand, and apply THRIVE principles in their daily work. This indicates that the NIP aids in embedding THRIVE principles within an organisation.

Previous research has found staff development and training are key to the successful implementation of a new intervention (Rahman et al., 2012; Resnick et al., 2018). Training activities and attendance at both i-THRIVE Academy and Community of Practice events were reported among implementation sites, and staff from implementation sites exhibited knowledge of THRIVE principles. Continued engagement of staff will likely be important for the integration of THRIVE, and formal assessments or competency checks may be useful to monitor THRIVE delivery (Sanders et al., 2022). In addition to educational activities, several of the implementation strategies used by the NIP, including site diagnostics activities, coaching and support, and involving key partners were found to have positive results at least 75% of the time in a recent review (Ashcraft et al., 2024)

Implementation research generates critical evidence for selecting effective strategies for health system transformation (Ashcraft et al., 2024). Like the NIP, most implementations include multiple strategies (Ashcraft et al., 2024), meaning it is difficult to attribute a successful implementation to any single strategy. There are few examples of implementation research for mental health, many of which focus on implementation effectiveness (Wells et al., 2013; Chung et al., 2014; Kilbourne et al., 2014; Waxmonsky et al., 2014; Chung et al., 2015; Sinnema et al., 2015; Morton et al., 2020; Ruud et al., 2021; Bartels et al., 2022; Toropova et al., 2022) with a minority examining the implementation process more broadly (Williams et al., 2017; Bauer et al., 2019; Leone et al., 2022). Among studies of the implementation process or implementation fidelity, a facilitator-implemented collaborative care model resulted in some improvements to team function; adoption of collaborative care model processes varied widely by site (Bauer et al., 2019). Another study examined mediating factors for clinician adoption of evidence-based practices following an organisation-supported implementation strategy, reporting high fidelity to the strategy at both clinician and organisation levels (Williams et al., 2017). Although implementation studies would benefit from enrolment of many participating sites to reduce the impact of between-site variation, the enrolment of a high number of sites presents many logistical challenges. Integrated care models, such as THRIVE, are used in Europe, Australia, and North America, and have been tailored for use in services for CYP (Hodgins et al., 2024). A review of integrated care models for youth discusses common features, including multidisciplinary staff members able to partner with external organisations and managers committed to integration, joint planning, and stakeholder partnership (Hodgins et al., 2024). The THRIVE Framework targets a local community, and relies on the involvement of multiple agencies to transform how these agencies provide mental healthcare for CYP (Wolpert et al.). Many of the implementation sites hosted events to build stakeholder relationships. In this study, we found that multi-agency cooperation was critical to the implementation of the NIP itself: the strength of working relationships in the local system moderated the effect of the NIP on THRIVE fidelity among sites. For future users of the NIP, an evaluation of local working relationships or pre-implementation efforts to engage with community stakeholders and strengthen these relationships would be worthwhile.

On average, the NIP had a modest impact on THRIVE fidelity among sites, without reaching the level of a statistically-significant change. With a small number of sites included, the study may have been under-powered. When implementation studies find a null effect, it is difficult to know if the effect is truly null or if the implementation itself was incorrect (Sanders et al., 2022), or if implementation success depends on site characteristics (Augustsson et al., 2015). Site-level changes in THRIVE fidelity had high variability; even when an implementation is supported in the same way for all sites, variation can occur (Augustsson et al., 2015). To better understand the impact of NIP, we examined the moderating effect of an important site characteristic: the strength of local working relationships at the site, finding that the NIP improved THRIVE fidelity (overall and at macro level) at implementation sites with strong local working relationships, compared to comparison sites with strong local working relationships. A similar result was found in a study of an organisational-level intervention for occupational health, where the implementation worked the best among units with strong collaboration (Augustsson et al., 2015). To explain these results, the authors suggest that the intervention was a better fit for those units or that the units were more capable in adapting the intervention. Preliminary work can be done to prepare sites for an implementation; for those sites interested in using the NIP, efforts to strengthen local working relationships prior to implementation should be considered.

The future direction of this research will involve comprehensive evaluations of the service and clinical impacts of the NIP. These evaluations will encompass a range of critical factors, including the accessibility and efficiency of services, clinical outcomes, patient experiences, and the specific impacts on various sub-groups within the patient population. The latter will particularly focus on racial or ethnic minorities and distinct diagnostic categories, ensuring a broad and inclusive understanding of the NIP’s effectiveness.

## Limitations

There are several limitations to consider in this work. For the survey of site staff, the response rate was low (28.5%) compared to staff surveys in other implementation studies (46.8—83.1%) (Leone et al., 2022; Toropova et al., 2022). We would expect this to cause some bias in our results if the response rate was differential by implementation/comparison site groups, but there was no difference in response rate by these groups. The low response rate may indicate that the survey results are not generalisable. Future research should consider incentives to encourage survey completion.

When introducing new health policies, understanding their impact and identifying the contexts in which they are most effective is crucial. Estimating the mean effect presents several methodological challenges, especially when data from pre-implementation periods are scarce or when only a few sites are involved. Unlike randomised controlled trials, the adoption of health policies is not random. This means that the characteristics of the implementation and control sites are likely to vary, leading to potential selection bias. Yet, it is a useful methodological approach for evaluating complex interventions selected and implanted by policymakers. It is often unclear which characteristics influence a site’s decision to adopt a health policy. Even with numerous characteristics measured, careful consideration of each characteristic’s role is necessary. Confounding is another potential bias, where some characteristics may influence both the decision to implement and the site’s capacity to do so effectively.

To adjust for these biases, various methods are available, but these methods can themselves introduce bias. Sensitivity analyses are essential in gaining a deeper understanding of the data. The four-group propensity-score weighting DiD method is designed to mitigate potential selection biases and confounding factors (Stuart et al., 2014). However, a key assumption of DiD, the parallel trends assumption, is not verifiable. Violations of the parallel trends assumption can lead to issues with time-varying confounding (Xu, 2017), complicating the interpretation of results. Thus, while our study provides valuable insights into the effectiveness of the NIP, these issues must be considered when interpreting the findings.

The variable transformation approaches used by comparison sites could be considered a limitation, as this could complicate the interpretation of results. Strictly speaking, a comparison group where all sites were transforming their CAMHS into the THRIVE Framework would allow us to test the implementation strategies of the NIP specifically (i.e. all sites seeking to fit the THRIVE Framework but the implementation group testing the NIP implementation strategies). Four comparison sites reported using THRIVE as their transformation model, but this sample size is too small for a full analysis. The comparison group simply represents routine implementation, a common approach for implementation research control groups (Smith and Hasan, 2020).

## Conclusions

This study’s investigation into the effectiveness of the NIP in England offers significant insights into the implementation of complex health interventions, particularly in the field of CAMHS. The findings underscore the importance of effective working relationships among local systems in the successful adoption and implementation of health policies like i-THRIVE. Specifically, the study demonstrates that sites with highly effective working relationships exhibit substantial improvements in adhering to THRIVE principles, as evidenced by the increase in fidelity scores.

The broad awareness of the THRIVE framework among staff, even in comparison sites, highlights the programme’s permeation in the field of CYPMH. However, implementation strategies are critical in embedding these principles more deeply within organisations. This distinction is crucial for policymakers and healthcare leaders aiming to foster more effective, integrated mental health services tailored to the needs of CYP.

Methodologically, the study navigates the challenges of evaluating health policy implementations in non-randomised settings. The use of four-group propensity-score weighted DiD analysis is an effective approach to address potential biases, such as selection bias and confounding factors. However, it also highlights the inherent complexities and limitations in evaluating policy impact in real-world settings. The study’s sensitivity analyses further strengthen the validity of its findings.

In conclusion, the NIP presents a promising model for CAMHS. Its emphasis on effective working relationships and the tailored approach to implementation are key factors in its success. The insights from this study contribute valuable knowledge to the ongoing efforts to improve mental health services and can guide future policies and programmes aimed at enhancing the well-being of CYP.

## Data Availability

All datasets and code are accessible on GitHub (Sippy, 2023).

https://zenodo.org/records/10418119

## Ethical Standards

The NHS/University Joint Research Office reviewed this study and classified it as a service evaluation, meaning it does not need ethical approval or a review by the Research Ethics Committee (IRAS application number: 250439). For the qualitative parts of the study, participants included implementation leads, managers, and front-line staff, who were interviewed about their service transformation projects. At the start of each interview, verbal informed consent was obtained, which included explicit permission for the interview to be recorded, analysed, and used in the service evaluation. This consent was recorded, and transcripts of each interview were made to document it.

The data for the quantitative evaluation is retrospective and was routinely collected from service records. It was anonymised at the source by the business intelligence staff at each trust, with all identifying information removed. After review by local information governance leads, this anonymised data was provided to the evaluation team. As the data is retrospective and de-identified, consent for its use in the evaluation is neither legally nor ethically required. The requirement for ethical approval was therefore waived by the joint research office.

## Availability of Data and Materials

All datasets and code are accessible on GitHub (Sippy, 2023).

## Acknowledgements

The authors are indebted to the clinicians, managers and academics who have created THRIVE and their generous help and thoughtful advice throughout the evaluation to assess its effectiveness, in particular Sophie Dunn. We would also like to thank Rudolf Cardinal for help with the research.

