## Supplemental Material S1 for "Effect of a needs-based model of care on the characteristics of healthcare services in England: the i-THRIVE National Implementation Programme"

**Supplemental Material S1: Delivery of i-THRIVE**

**National i-THRIVE Programme**

The implementation mode for i-THRIVE l is fully described in the study protocol (Moore et al., 2023). Briefly, the National i-THRIVE Programme translates the macro-, meso-, and micro-level principles of THRIVE into six components to aid the implementation process. Brief descriptions of each component are in Table S1. Sites use the i-THRIVE Approach to Implementation, which has four phases, described in Table S2.

| Table S1 National i-THRIVE Programme Components | |
| --- | --- |
| i-THRIVE Component | **Description** |
| *i-THRIVE Community of Practice* | All implementing sites can participate in shared learning events, where active sites can share their implementation process and support one another |
| *i-THRIVE Implementation Support Team* | This team oversees the i-THRIVE Programme, Community of Practice, and Academy, and developed the Toolkit. Local services receive focused support from the team |
| *i-THRIVE Approach to Implementation* | This is a manual describing the four-phase, structured approach with definitions for integration and recommended evaluations |
| *i-THRIVE Toolkit* | This is a resource to support sites in fidelity to the implementation protocol but is flexible to the needs of the local context |
| *i-THRIVE Academy* | This is a series of coaching and training in the THRIVE principles targeting the frontline staff |
| *i-THRIVE Option Grids* | These are aids for decision-making to help staff, families, and CYP have productive conversations regarding care |

| Table S2 Phases of i-THRIVE Approach to Implementation | |
| --- | --- |
| Phase | **Description** |
| *One* | Comprehensive understanding of local systems and population needs |
| *Two* | Identifying training needs of staff and building system capacity |
| *Three* | Implementing the new system and establishing information infrastructures |
| *Four* | Learning, embedding, and sustaining changes to the system |

**Implementation Site Progress**

Annual progress reports were published in May 2017 and May 2018, and included updates from the implementation sites (National i-THRIVE Programme Team, 2017). In addition, the i-THRIVE website includes summaries for each implementation site (i-THRIVE Team). Key achievements for each site are summarised in Table S3.

| Table S3: i-THRIVE Progress | |
| --- | --- |
| Site | **Achievements** |
| Site B | - Hosted a system-wide engagement event with over 100 participants - Mapped the needs and demands of the area - Collaboration with partner agencies and stakeholders |
| Site C | - Completed an in-depth analysis of system services - Developed an app fo children and young people to better communicate with their clinicians and follow their progress - Offered additional support for those with long-term health conditions - Hosted a THRIVE clinic |
| Site G | - Established an integrated partnership to focus on i-THRIVE - Mapped the care pathways in Stockport - Established a group to focus on mental health and emotional wellbeing in schools |
| Site L | - Engaged with third-sector organisations to employ THRIVE practitioners supporting CAMHS - Developed integrated access and care pathways - Staff attended i-THRIVE Academy - Used a radio programme to support children and young people - Offered additional support for those with eating disorders |
| Sites D & M | - Programme of staff training events - Clinician attendance at i-THRIVE Academy - THRIVE needs-based groupings included in case records - Developed an in-school resilience programme |
| Site O | - Focused on engagement with schools - Hosted a multi-agency engagement event with the i-THRIVE Implementation Support Team - Developed multi-disciplinary early help teams to support children and young people |
| Site Q | - Implementation of multi-agency local agreements for collective accountability - Integration of THRIVE needs-based grouping into the clinical record system - Provided services in schools - Integrated THRIVE needs-based groupings into care plans |
| Site R | - Developed a website for children, young people, and their families - Adoption of THRIVE by the local authority - Full review of CAMHS system with the i-THRIVE Implementation Support Team |
| Site T | - Developed a website and services tailored to local children and young people - Clinicians trained in shared decision making (i-THRIVE Academy) |

**Implementation Leads Self-Reported Experience**

As reported in the main manuscript, eight programme managers from seven implementation sites provided feedback on their site’s experience using i-THRIVE as part of the survey. The transformation leads survey responses were from NIP sites (Sites D, G, L (two managers), M, O, Q, and R). Responses related the planning of i-THRIVE are reported in Supplementary Figures 1 & 2. Responses related to the delivery of i-THRIVE are reported in Supplementary Figures 3 & 4.

**Supplementary Figure 1** Planning of i-THRIVE

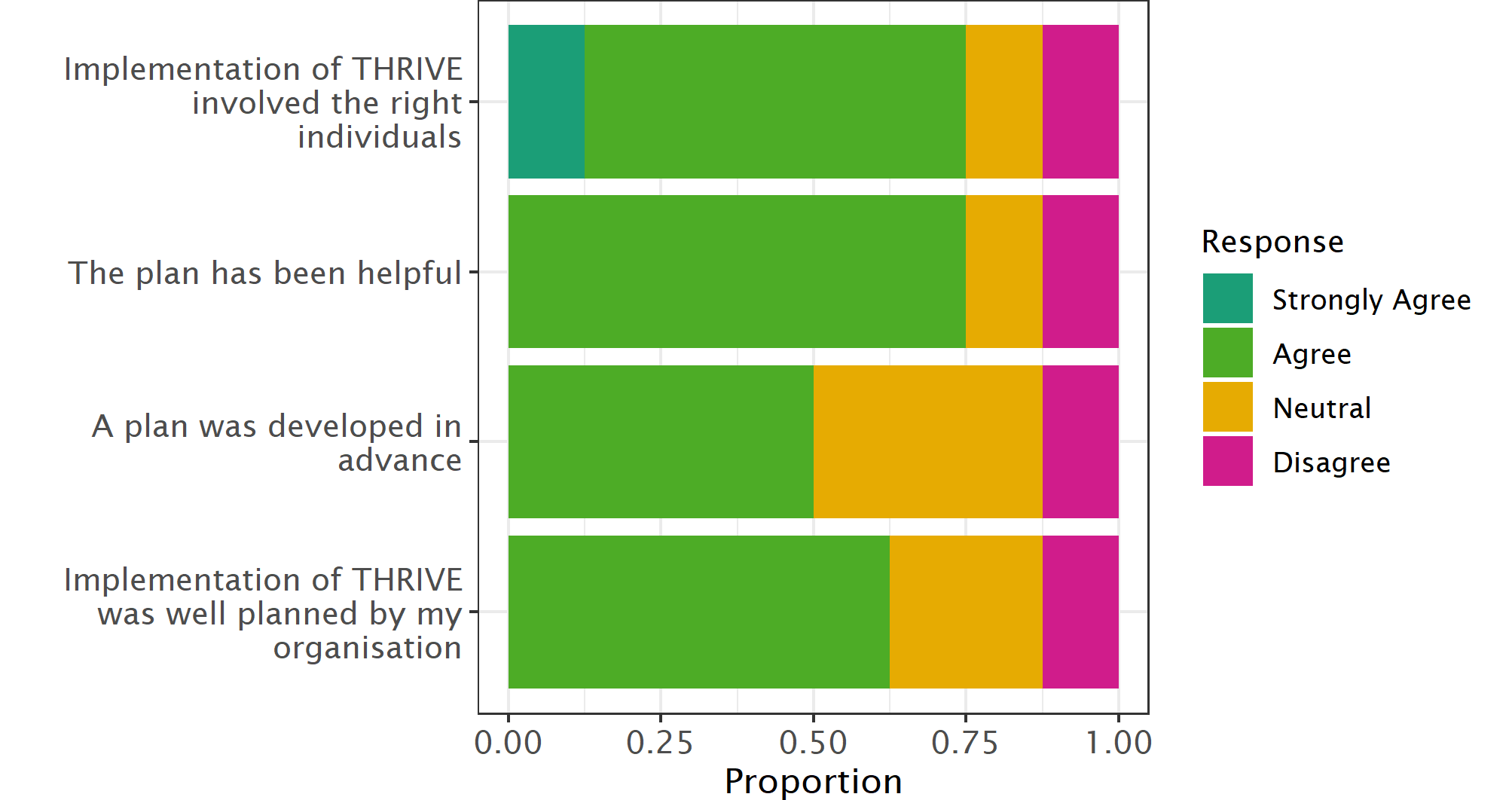

**Supplementary Figure 2** Individuals Involved in i-THRIVE Planning

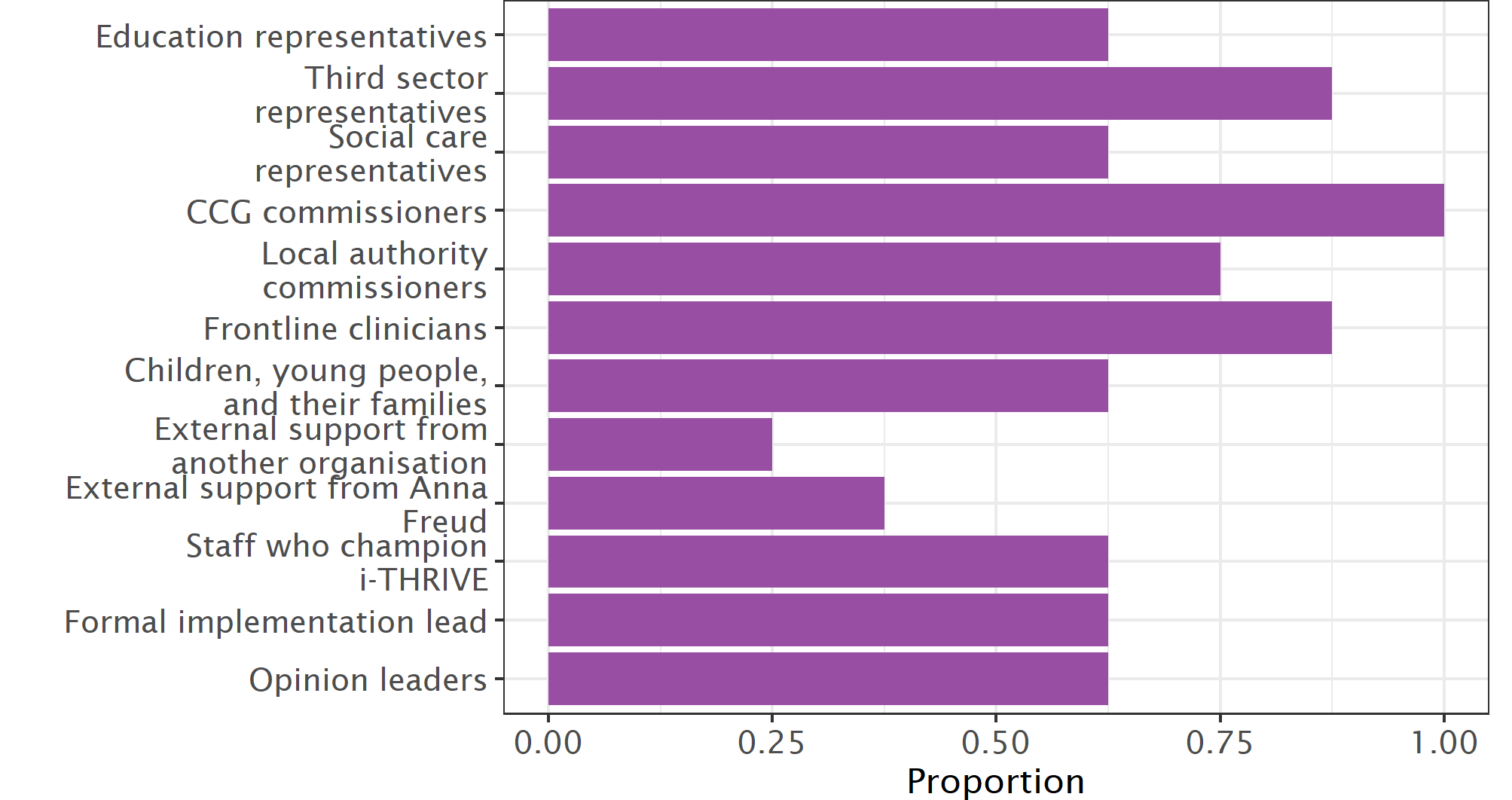

**Supplementary Figure 3** Programme Managers Perspectives of i-THRIVE Delivery

There were two non-responses to the third and fourth questions**.**

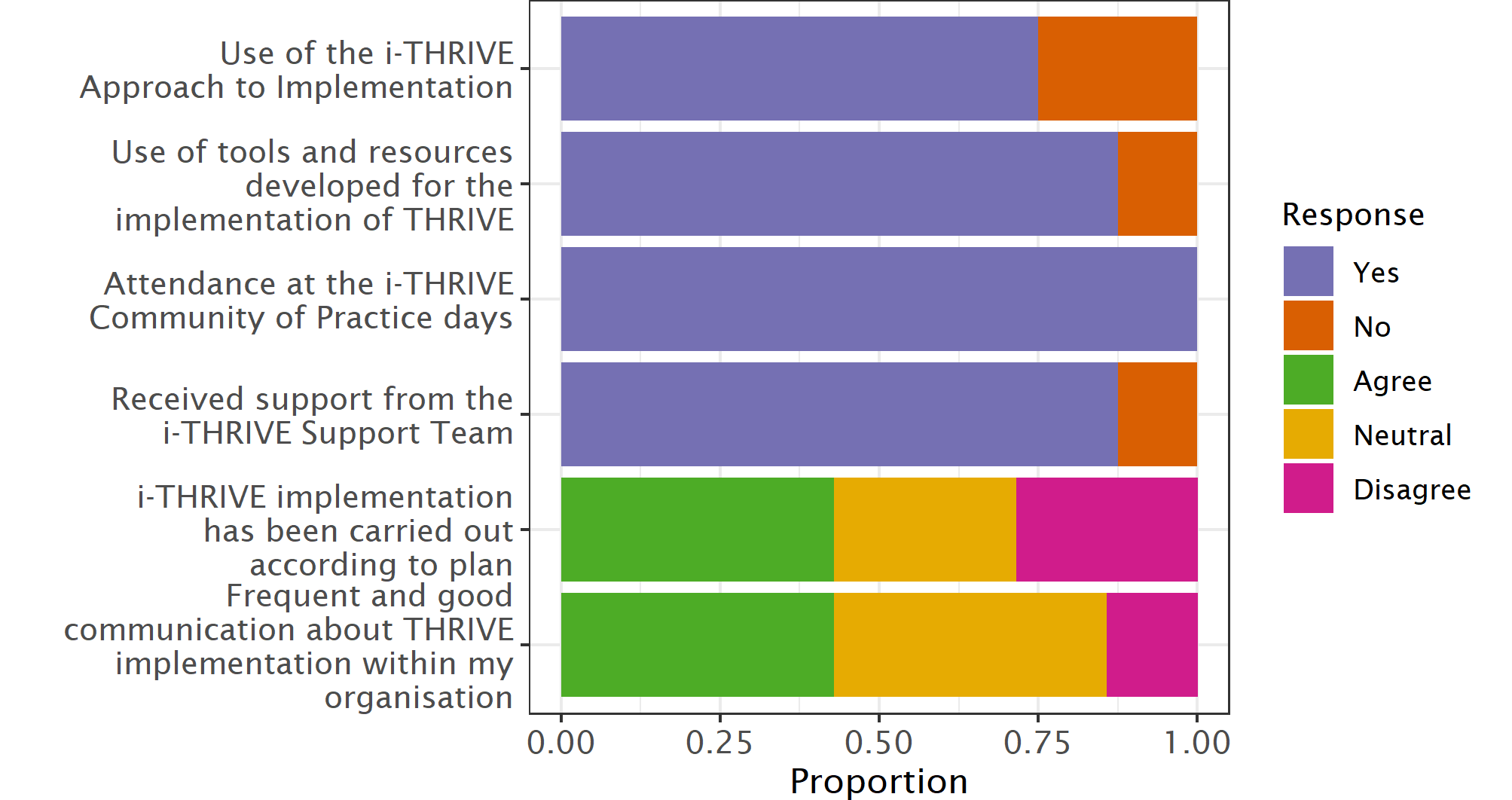

**Supplementary Figure S4** Individuals Involved in Implementation

One site (R) did not respond to this series of questions, so the proportion is calculated from seven sites. There was also one non-response for the first and fourth questions.

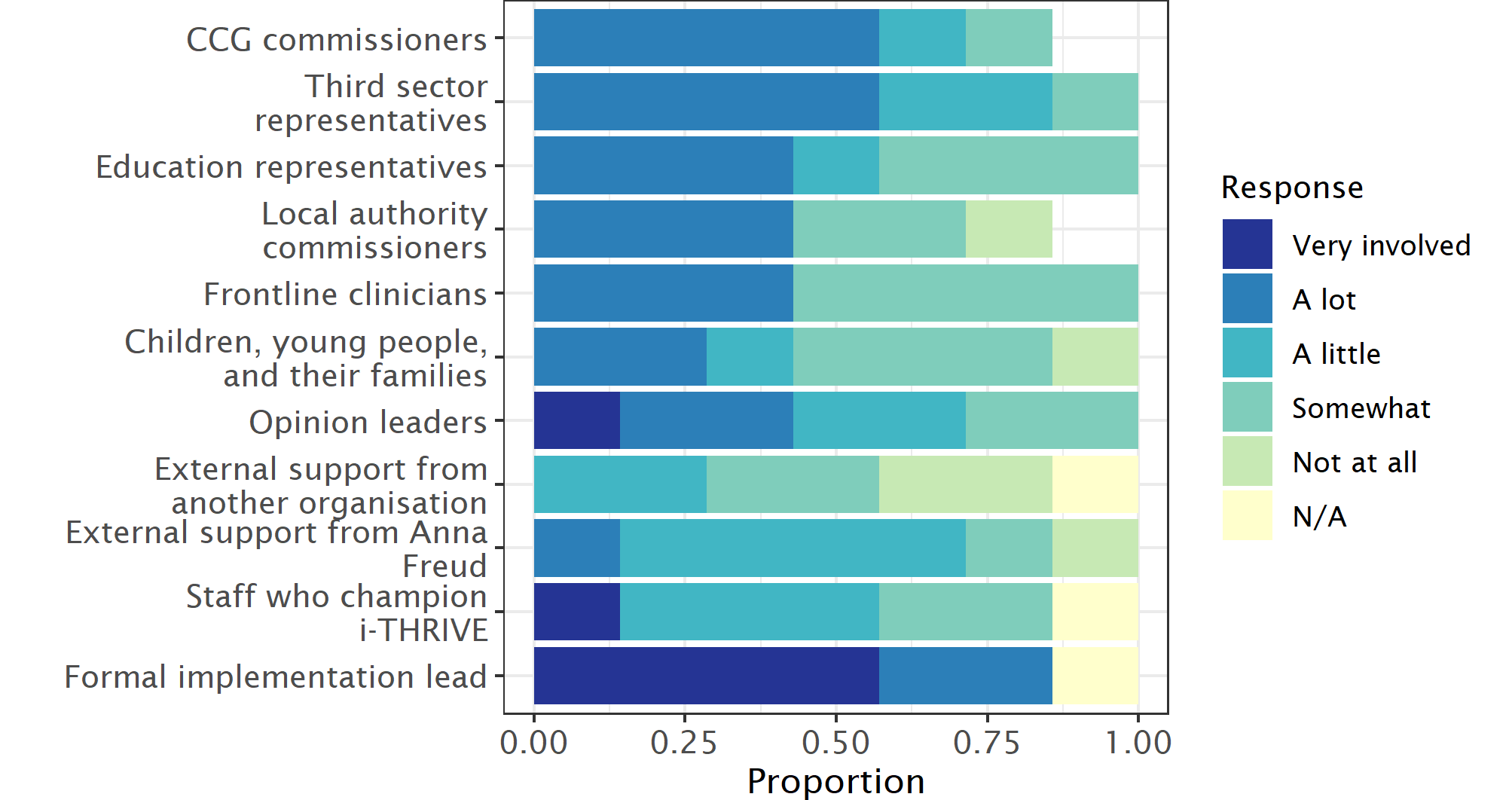

Most programme managers agreed that i-THRIVE was well-planned by their organisations, that the planning was helpful, and that the planning process involved the right individuals. Only half of the managers reported that the plan had been developed in advance. Many individuals were involved in the planning process, with most programme managers reporting planning involvement by those within the organisation at multiple levels (leaders and staff), individuals from other agencies (education, social care, local authorities, CCGs, and third sector), and importantly, five sites included children, young people, and their families in the planning process. There was less reliance on individuals from external organisations for planning.

Most programme managers reported used of the i-THRIVE Approach to Implementation (all except Sites M and Q), using tools and resources (all except Site G), and receiving support from the i-THRIVE Support Team (all except Site Q). All programme managers reported attendance at the i-THRIVE Community of Practice events. Two programme managers disagreed with the statement “*i-THRIVE implementation has been carried out according to plan*” (Sites G and L). There were two programme managers surveyed from Site L; the other programme manager for this site agreed with the statement. The Site G programme manager also disagreed with the statement “*There is frequent and good communication about how THRIVE implementation is going within my organisation*”. More than half of programme managers reported that formal implementation leads, third sector representatives, and CCG commissioners were “very involved” or had “a lot” of involvement in implementation.
