## Supplemental Material S2 for "Effect of a needs-based model of care on the characteristics of healthcare services in England: the i-THRIVE National Implementation Programme"

| Table S1 The TIDieR statement – checklist of items to include when describing an intervention and the location of the information | | | | |
| --- | --- | --- | --- | --- |
| Topic | **Item number** | **TIDieR items** | **Location where items are reported** | |
|  |  |  | Manuscript | Other |
| Brief name | 1 | Provide the name or a phrase that describes the intervention. | 1 | - |
| Why | 2 | Describe any rationale, theory, or goal of the elements essential to the intervention. | 4, 5 | - |
| What | 3 | Materials: Describe any physical or informational materials used in the intervention, including those provided to participants or used in intervention delivery or in training of intervention providers. Provide information on where the materials can be accessed (e.g. online appendix, URL). | - | (Moore et al., 2023) |
|  | 4 | Procedures: Describe each of the procedures, activities, and/or processes used in the intervention, including any enabling or support activities. | - | (Moore et al., 2023) |
| Who provided | 5 | For each category of intervention provider (e.g. psychologist, nursing assistant), describe their expertise, background and any specific training given. | - | (Moore et al., 2023) |
| How | 6 | Describe the modes of delivery (e.g. face-to-face or by some other mechanism, such as internet or telephone) of the intervention and whether it was provided individually or in a group. | - | (Moore et al., 2023) |
| Where | 7 | Describe the type(s) of location(s) where the intervention occurred, including any necessary infrastructure or relevant features. | 5, 6, Supplemental Material S3, Supplemental Material S7 | (Moore et al., 2023) |
| When and how much | 8 | Describe the number of times the intervention was delivered and over what period of time including the number of sessions, their schedule, and their duration, intensity or dose. |  | (Moore et al., 2023) |
| Tailoring | 9 | If the intervention was planned to be personalised, titrated or adapted, then describe what, why, when, and how. | - | (Moore et al., 2023) |
| Modifications | 10 | If the intervention was modified during the course of the study, describe the changes (what, why, when, and how). | - | (Moore et al., 2023) |
| How well | 11 | Planned: If intervention adherence or fidelity was assessed, describe how and by whom, and if any strategies were used to maintain or improve fidelity, describe them. | - | (Moore et al., 2023) |
|  | 12 | Actual: If intervention adherence or fidelity was assessed, describe the extent to which the intervention was delivered as planned. | Supplemental Material S1 | - |

** **Authors** - use N/A if an item is not applicable for the intervention being described. **Reviewers** – use ‘?’ if information about the element is not reported/not sufficiently reported.
