## Supplemental Material S3 for "Effect of a needs-based model of care on the characteristics of healthcare services in England: the i-THRIVE National Implementation Programme"

**Supplemental Material S3: Additional Methodological Details**

This section of the supplemental materials provides further details of the methods used for our study.

**Study Setting and Design**

Most sites were equivalent to a single CCG, but five included multiple CCGs (see Table S1). There were administrative changes during the study in April 2017: Central Manchester, North Manchester, and South Manchester CCGs merged to form the Manchester CCG, eight GP practices moved from South Norfolk CCG to Norwich CCG, and one GP practice transitioned from Bradford City to Bradford Districts (NHS England, 2019).

| **Sites** | | **CCGs** |
| --- | --- | --- |
| i-THRIVE | Bexley | - |
|  | Cambridgeshire and Peterborough | - |
|  | Camden | - |
|  | Hertfordshire | 1. East and North Hertfordshire 2. Herts Valley |
|  | Luton | - |
|  | Manchester | 1. Central Manchester 2. North Manchester 3. Salford 4. South Manchester |
|  | Stockport | - |
|  | Tower Hamlets | - |
|  | Waltham Forest | - |
|  | Warrington | - |
| Comparison | Bradford | 1. Airedale, Wharfdale and Craven 2. Bradford City 3. Bradford Districts |
|  | Ipswich and East Suffolk | - |
|  | Lewisham | - |
|  | Norfolk | 1. Great Yarmouth and Waveney 2. North Norfolk 3. Norwich 4. South Norfolk 5. West Norfolk |
|  | Northampton | 1. Corby 2. Nene |
|  | Portsmouth | - |
|  | Southampton | - |
|  | Stoke-on-Trent | - |
|  | Sunderland | - |
|  | South Worcestershire | - |

**Surveys**

In the staff survey, staff were asked to identify five THRIVE needs categories correctly, to evaluate knowledge of the THRIVE Framework. As part of a validation process, fifty individuals not involved in the evaluation took this assessment. Those familiar with THRIVE showed a 75% or higher accuracy rate, while those unfamiliar scored 40% or lower. This result suggests the questions effectively measured specific knowledge related to the programme rather than general tendencies or biases.

The transformation leads survey responses were from NIP sites (Sites D, G, L (two managers), M, O, Q and R) as well as comparison sites (Sites A, E, I, Bradford, EastSuff, J (two managers), K, P, and S).

**Fidelity Ratings**

Fidelity scoring was based on information collected during interviews using the i-THRIVE Assessment Tool and quantitative data on service use collected from the NHS. Interviewees were drawn from multiple agencies, including CAMHS, CCGs, education, local authorities, and third sector. All twenty sites received a fidelity score for 75 THRIVE principles, before (between December 2017 and July 2019) and after implementation (between October 2018 and February 2020), resulting in 40 sets of scores. The median interval period between baseline and follow-up scoring was 178 days. Interviews were conducted over the phone and were recorded, lasting from 45 minutes to 1 hour. A total of 352 interviews were conducted (182 baseline interviews and 170 follow-up interviews). At least two independent raters scored for each set. To determine the interrater reliability among the raters of a set, Krippendorff’s alpha was calculated (Hayes and Krippendorff, 2007). This metric ranges from 0 (low) to 1 (high) and is appropriate for two or more raters providing ordinal scores. The i-THRIVE Assessment Tool has not been validated.
