## Supplemental Material S4 for "Effect of a needs-based model of care on the characteristics of healthcare services in England: the i-THRIVE National Implementation Programme"

**Supplemental Material S4: Survey for CAMHS Staff**

1. Which of the following best describes your organisation?
   a. NHS
   b. CCG
   c. Local authority
   d. Third sector
   e. Education
   f. Other (please state)
2. Which of the following best describes the primary purpose of the service that you work in?
   a. Specialist mental health services
   b. Brief intervention/counselling or support
   c. A service that is aimed at preventing mental health problems in children
   d. Supporting children and young people in the community who are considered to be high risk
   e. Triage, signposting, advice or assessment
   f. Children’s community services
   g. Social care
   h. Education
   i. Commissioning
   j. Admin
   k. Other (please state)
3. Which of the following best describes how you spend the majority of your time in your current role?
   a. Frontline practitioner with direct contact with children and young people
   b. Manager of a service or team
   c. Senior leadership within an organisation
   d. Commissioner
   e. Admin
   f. Other (please state)
4. Have you heard of the THRIVE Framework?
   a. Yes
   b. No
5. How have you heard of THRIVE? Please select all that apply.
   a. Communications from within my organisation (eg. newsletter, website, other communications to staff)
   b. Communications from outside my organisation (eg. local policy, reports from other organisations)
   c. Discussed during a training course, event or meeting
   d. In the media or online
   e. Talking to colleagues
   f. Talking to friends or family (not including colleagues)
6. Do you consider your service to be implementing THRIVE?
   a. Yes
   b. No
   c. Unsure
   d. (Optional) If you are unsure, could you provide some more information?
7. Do you personally feel that you use THRIVE principles in your daily practice?
   a. Yes
   b. No
   c. Unsure
   d. (Optional) If so, in what ways do you use THRIVE in your daily practice?
8. To your knowledge, is there a quality improvement initiative focused on implementing THRIVE in your organisation? This could be in a different service than the one where you work.
   a. Yes
   b. No
   c. (Optional) Do you know what aspect of the service the QI initiative is focused on improving?
9. Which of the following are THRIVE groups? Please select all that apply.
   a. Getting advice and signposting
   b. Getting more help
   c. Getting crisis support
   d. Getting help
   e. Getting risk support
   f. Getting specialist services
