## Supplemental Material S5 for "Effect of a needs-based model of care on the characteristics of healthcare services in England: the i-THRIVE National Implementation Programme"

### i-THRIVE Survey - Implementation Leads

#### Five minute survey - we would really appreciate your help!

**We'd like to hear your views about the implementation of the THRIVE Framework in your area. Your responses are important for us to understand the factors that contribute to its implementation.**

**We thank you for your help!**

**For any questions, please contact Liz Simes, Research Manager, at. More information is available on the i-THRIVE website.**

### i-THRIVE Survey - Implementation Leads

#### Questions about yourself

1. Which of the following best describes your organisation?

- NHS
- CCG
- Local Authority
- Third Sector
- Education
- Other (please state)

2. Please select the locality you work in.

3. Which of the following best describes the primary purpose of the service that you work in?

- Specialist mental health services
- Brief intervention / counselling or support
- A service that is primarily aimed at preventing mental health problems in children
- Supporting children and young people in the community who are considered to be high risk
- Triage, signposting, advice or assessment
- Other (please state)

4. Please provide the name of your organisation.

5. Which of the following best describes how you spend the majority of your time in your current role?

- ○ Frontline practitioner with direct contact with children and young people
- ○ Manager of a service or team
- ○ Senior leadership within an organisation
- ○ Commisioner
- ○ Other (please state)

6. How long have you been employed by your organisation?

- ○ Less than 1 year
- ○ 1 - 5 years
- ○ 5 - 10 years
- ○ 10 - 15 years
- ○ 15 - 20 years
- ○ Over 20 years

7. Have you been involved in the implementation of THRIVE in your locality?

- ○ Yes
- ○ No

#### Developing a plan to implement THRIVE

8. Is your organisation implementing THRIVE?

- ◯ Yes
- ◯ No
- ◯ To some extent

If you have ticked ’to some extent’ would you please provide a brief explanation of this answer.

9. Please select the extent to which you agree with the following statements around planning implementation.

| | Strongly agree | Agree | Neither agree nor disagree | Disagree | Strongly disagree |
|---|---|---|---|---|---|
| A plan for implementing THRIVE was developed in advance | ◯ | ◯ | ◯ | ◯ | ◯ |
| The implementation plan has been helpful | ◯ | ◯ | ◯ | ◯ | ◯ |
| Implementation of THRIVE involved the right individuals | ◯ | ◯ | ◯ | ◯ | ◯ |
| Planning for the implementation of THRIVE was well done by my organisation | ◯ | ◯ | ◯ | ◯ | ◯ |

10. Who was involved in developing a plan to implement THRIVE? Please select all that apply.

- [ ] Opinion leaders - active and respected voices within the service
- [ ] Formally appointed i-THRIVE implementation lead
- [ ] Staff who champion i-THRIVE in addition to their regular duties
- [ ] External implementation support from the Anna Freud Centre
- [ ] External implementation support from another external organisation (eg. NHS Improvement)
- [ ] Children, young people and their families
- [ ] Frontline clinicians
- [ ] Commissioners from the local authority
- [ ] Commissioners from the CCG
- [ ] Representatives from social care
- [ ] Representatives from the third sector
- [ ] Representatives from education

Other (please specify)

#### Implementation tools to guiding local i-THRIVE approach

11. Did you use the "i-THRIVE Approach to Implementation" to help guide your approach?*

* The "i-THRIVE Apporach to Implementation" has been developed by the Anna Freud Centre and the Tavistock and Portman NHS Foundation Trust and is designed to guide services through the process of implementing THRIVE.

- ◯ Yes
- ◯ No
- ◯ I don't know
- ◯ To some extent

If you have ticked 'to some extent' would you please provide a brief explanation of this answer.

12. Please answer the following questions about your use of the implementation support by the National i-THRIVE implementation Team based at the Anna Freud Centre and the Tavistock and Portman NHS Foundation Trust.

| | Yes | No |
|---|---|---|
| Did you use the "i-THRIVE Approach to Implementation" to help guide your approach? | ◯ | ◯ |
| Did you use any of the implementation tools developed for i-THRIVE implementation (available online or through the i-THRIVE Programme?) | ◯ | ◯ |
| Did you attend any of the i-THRIVE Community of Practice days? | ◯ | ◯ |
| Did you receive support from the i-THRIVE Implementation Support Team at the Anna Freud Centre or the Tavistock and Portsman NHS Foundation Trust? | ◯ | ◯ |

13. Please add any further comments about the i-THRIVE Implementation Programme.

14. Did you use any other implementation approaches to help guide your approach?

- ◯ Yes
- ◯ No

15. If yes, was this a structured approach to implementation?

- ◯ Yes
- ◯ No

If yes, please provide further details

#### Involving others in implementing THRIVE

16. To what extent do you agree with the following statements?

| | Strongly agree | Agree | Neither agree nor disagree | Disagree | Strongly disagree |
|---|---|---|---|---|---|
| Some of our staff have become i-THRIVE champions, actively supporting and promoting THRIVE implementation beyond what is required | ○ | ○ | ○ | ○ | ○ |
| Clinical staff take an active interest in i-THRIVE related problems and successes | ○ | ○ | ○ | ○ | ○ |
| Managers in the service actively support i-THRIVE implementation | ○ | ○ | ○ | ○ | ○ |
| Commissioners actively support the implementation of i-THRIVE | ○ | ○ | ○ | ○ | ○ |
| Other agencies in the local system actively support i-THRIVE implementation | ○ | ○ | ○ | ○ | ○ |

17. To what extent were the following people involved in implementing THRIVE?

| | Very involved | A lot | Somewhat involved | A little | Not at all involved | N/A (Role does not exist) |
|---|---|---|---|---|---|---|
| Formally appointed i-THRIVE implementation lead | ◯ | ◯ | ◯ | ◯ | ◯ | ◯ |
| Staff who champion i-THRIVE in addition to their regular duties | ◯ | ◯ | ◯ | ◯ | ◯ | ◯ |
| External implementation support from the Anna Freud Centre or the Tavistock and Portman NHS Foundation Trust | ◯ | ◯ | ◯ | ◯ | ◯ | ◯ |
| External implementation support from another organisation (eg. NHS Improvement) | ◯ | ◯ | ◯ | ◯ | ◯ | ◯ |
| Opinion leaders - active and respected voices in the service | ◯ | ◯ | ◯ | ◯ | ◯ | ◯ |
| Children, young people and their families | ◯ | ◯ | ◯ | ◯ | ◯ | ◯ |
| Frontline clinicians | ◯ | ◯ | ◯ | ◯ | ◯ | ◯ |
| Local authority or social care representatives | ◯ | ◯ | ◯ | ◯ | ◯ | ◯ |
| Education representatives | ◯ | ◯ | ◯ | ◯ | ◯ | ◯ |
| Third sector representatives | ◯ | ◯ | ◯ | ◯ | ◯ | ◯ |
| CCG representatives | ◯ | ◯ | ◯ | ◯ | ◯ | ◯ |

18. Do you feel the correct people were involved in carrying out implementation? If not, who should have been?

19. To what extent to you agree with the following statements?

| | Strongly agree | Agree | Neither agree or disagree | Disagree | Strongly disagree |
|---|---|---|---|---|---|
| My organisation works well with the local authority to meet children and young people's mental health needs in my area | ○ | ○ | ○ | ○ | ○ |
| My organisation works well with schools to meet children and young people's mental health needs in my area | ○ | ○ | ○ | ○ | ○ |
| My organisation works well with the voluntary sector to meet children and young people's mental health needs in my area | ○ | ○ | ○ | ○ | ○ |
| My organisation works well with the NHS to meet children and young people's mental health needs in my area | ○ | ○ | ○ | ○ | ○ |
| Health and social care commissioners work well together in my area | ○ | ○ | ○ | ○ | ○ |
| My organisation generally has good relationships with other agencies in my area | ○ | ○ | ○ | ○ | ○ |
| The right organisations were involved in the implementation of THRIVE | ○ | ○ | ○ | ○ | ○ |

(Optional) Please provide any further comments

### i-THRIVE Survey - Implementation Leads

#### Reflecting on the process of implementation

20. To what extent do you agree with the following statements?

| | Strongly agree | Agree | Neither agree nor disagree | Disagree | Strongly disagree |
|---|---|---|---|---|---|
| i-THRIVE implementation has been carried out according to plan to date | ◯ | ◯ | ◯ | ◯ | ◯ |
| There is frequent and good communication about how THRIVE implementation is going within my organisation | ◯ | ◯ | ◯ | ◯ | ◯ |
| Data is used to guide i-THRIVE implementation within my organisation | ◯ | ◯ | ◯ | ◯ | ◯ |

(Optional) Please provide any comments about how the implementation process has gone

21. Are you happy to be contacted for further details about this survey? If yes, please add your email below.
