## Supplemental Material S6 for "Effect of a needs-based model of care on the characteristics of healthcare services in England: the i-THRIVE National Implementation Programme"

### i-THRIVE Questionnaire - CAMHS Transformation Leads

Five minute survey - we would really appreciate your help!

**We'd like to hear your views about the transformation on the mental health services for children and young people in your area. Your responses are important for us to understand the factors that can impact the CAMHS re-design locally.**

**We thank you for your help!**

**For any questions, please contact Liz Simes, Research Manager, at. More information is available on the i-THRIVE website.**

#### i-THRIVE Questionnaire - CAMHS Transformation Leads

##### Questions about yourself

1. Which of the following best describes your organisation?

- ◯ NHS
- ◯ CCG
- ◯ Local Authority
- ◯ Third Sector
- ◯ Education
- ◯ Other (please state)

2. Please select the locality you work in.

3. Which of the following best describes the primary purpose of the service that you work in?

- ◯ Specialist mental health services
- ◯ Brief intervention / counselling or support
- ◯ A service that is primarily aimed at preventing mental health problems in children
- ◯ Supporting children and young people in the community who are considered to be high risk
- ◯ Triage, signposting, advice or assessment
- ◯ Other (please state)

4. Please provide the name of your organisation.

5. Which of the following best describes how you spend the majority of your time in your current role?

- ◯ Frontline practitioner with direct contact with children and young people
- ◯ Manager of a service or team
- ◯ Senior leadership within an organisation
- ◯ Commisioner
- ◯ Other (please state)

6. How long have you been employed by your organisation?

- ◯ Less than 1 year
- ◯ 1 - 5 years
- ◯ 5 - 10 years
- ◯ 10 - 15 years
- ◯ 15 - 20 years
- ◯ Over 20 years

7. Have you been involved in the CAMHS transformation in your locality?

- ◯ Yes
- ◯ No

#### i-THRIVE Questionnaire - CAMHS Transformation Leads

##### Developing a plan for CAMHS transformation

8. Does your organisation implement the CAMHS trasformation plan developed by local commissioners?

- Yes
- No
- To some extent

If you have ticked ’to some extent’ would you please provide a brief explanation of this answer.

9. Please select the extent to which you agree with the following statements around planning implementation.

| | Strongly agree | Agree | Neither agree nor disagree | Disagree | Strongly disagree |
|---|---|---|---|---|---|
| The implementation of CAMHS transformation was well planned by my organisation | | | | | |
| A plan for implementing CAMHS transformation was developed in advance | | | | | |
| The implementation plan has been helpful | | | | | |
| Implementing CAMHS transformation involved the right individuals | | | | | |

10. Who was involved in developing a plan for transforming CAMHS? Please select all that apply.

- [ ] Opinion leaders - active and respected voices within the service
- [ ] Formally appointed CAMHS transformation lead
- [ ] Staff who champion CAMHS transformation in addition to their regular duties
- [ ] Implementation support from an extneral organisation (e.g. research centre, university, NHS Improvement)
- [ ] Children, young people and their families
- [ ] Frontline clinicians
- [ ] Commissioners from the local authority
- [ ] Commissioners from the CCG
- [ ] Representatives from social care
- [ ] Representatives from the third sector
- [ ] Representatives from education

Other (please specify)

#### Implementation tools to guiding local CAMHS Transformation

11. Did you use a service model or approach to help you guide CAMHS transformation (e.g. CAPA, the THRIVE Framework)?

- Yes
- No

12. Did you use tools or resources to help you guide local CAMHS Transformation?

- Yes
- No
- I don't know
- To some extent

If you have ticked ’to some extent’ would you please provide a brief explanation of this answer.

13. Did you receive support from external organisations (e.g. research centre, university, NHS Improvement) to help you guide local CAMHS transformation?

- Yes
- No
- To some extent

If you have ticked ’to some extent’ would you please provide a brief explanation of this answer.

14. Did you attend any events that helped you with local CAMHS transformation?

- Yes
- No

If yes, would you please provide more details to you answer.

#### i-THRIVE Questionnaire - CAMHS Transformation Leads

##### Involving others in transforming CAMHS

15. To what extent do you agree with the following statements?

| | Strongly agree | Agree | Neither agree nor disagree | Disagree | Strongly disagree |
|---|---|---|---|---|---|
| Some of our staff have become champions for implementing CAMHS transformation, actively supporting and promoting it beyond what is required | ○ | ○ | ○ | ○ | ○ |
| Clinical staff take an active interest in the CAMHS transformation related problems and successes | ○ | ○ | ○ | ○ | ○ |
| Managers in the service actively support the implementation of the CAMHS transformation | ○ | ○ | ○ | ○ | ○ |
| Commissioners actively support the implementation of CAMHS transformation | ○ | ○ | ○ | ○ | ○ |
| Other agencies in the local system actively support the implementation of CAMHS transformation | ○ | ○ | ○ | ○ | ○ |

16. To what extent were the following people involved in implementing CAMHS transformation?

| | Very involved | A lot | Somewhat involved | A little | Not at all involved | N/A (Role does not exist) |
|---|---|---|---|---|---|---|
| Formally appointed CAMHS transformation lead | ○ | ○ | ○ | ○ | ○ | ○ |
| Staff who champion CAMHS transformation in addition to their regular duties | ○ | ○ | ○ | ○ | ○ | ○ |
| External implementation support from expert organisations (eg. NHS Improvement, Anna Freud Centre) | ○ | ○ | ○ | ○ | ○ | ○ |
| Opinion leaders - active and respected voices in the service | ○ | ○ | ○ | ○ | ○ | ○ |
| Children, young people and their families | ○ | ○ | ○ | ○ | ○ | ○ |
| Frontline clinicians | ○ | ○ | ○ | ○ | ○ | ○ |
| Local authority or social care representatives | ○ | ○ | ○ | ○ | ○ | ○ |
| Education representatives | ○ | ○ | ○ | ○ | ○ | ○ |
| Third sector representatives | ○ | ○ | ○ | ○ | ○ | ○ |
| CCG representatives | ○ | ○ | ○ | ○ | ○ | ○ |

17. Do you feel the correct people were involved in carrying out implementation? If not, who should have been?

18. To what extent to you agree with the following statements?

| | Strongly agree | Agree | Neither agree or disagree | Disagree | Strongly disagree |
|---|---|---|---|---|---|
| My organisation works well with the local authority to meet children and young people's mental health needs in my area | ○ | ○ | ○ | ○ | ○ |
| My organisation works well with schools to meet children and young people's mental health needs in my area | ○ | ○ | ○ | ○ | ○ |
| My organisation works well with the voluntary sector to meet children and young people's mental health needs in my area | ○ | ○ | ○ | ○ | ○ |
| My organisation works well with the NHS to meet children and young people's mental health needs in my area | ○ | ○ | ○ | ○ | ○ |
| Health and social care commissioners work well together in my area | ○ | ○ | ○ | ○ | ○ |
| My organisation generally has good relationships with other agencies in my area | ○ | ○ | ○ | ○ | ○ |
| The right organisations were involved in the implementation of CAMHS Transformation | ○ | ○ | ○ | ○ | ○ |

(Optional) Please provide any further comments

#### i-THRIVE Questionnaire - CAMHS Transformation Leads

##### Reflecting on the process of implementation

19. To what extent do you agree with the following statements?

| | Strongly agree | Agree | Neither agree nor disagree | Disagree | Strongly disagree |
|---|---|---|---|---|---|
| The implementation of CAMHS transformation has been carried out according to plan to date | ◯ | ◯ | ◯ | ◯ | ◯ |
| There is frequent and good communication about how the implementation of CAMHS transformation is going within my organisation | ◯ | ◯ | ◯ | ◯ | ◯ |
| Data is used to guide the implementation of CAMHS transformation within my organisation | ◯ | ◯ | ◯ | ◯ | ◯ |

(Optional) Please provide any comments about how the implementation process has gone

[ ]

20. Are you happy to be contacted for further details about this survey? If yes, please add your email below.

[ ]
