## Supplemental Material S7 for "Effect of a needs-based model of care on the characteristics of healthcare services in England: the i-THRIVE National Implementation Programme"

**Supplemental Material S7: Sensitivity Analyses**

This section of the supplemental materials provides a detailed description of the sensitivity analyses conducted for our study. These analyses are crucial in validating the robustness of our findings and include the use of alternative methods for health policy analysis, different model specifications for effect estimation, and the consideration of excluding non-compliant control sites. Additionally, we present the results of these analyses together, facilitating a straightforward comparison.

**Alternative Method of Analysis**

For the estimation of the NIP effect, we employed two alternative methods: the standard difference in differences (DiD) and the four-group propensity score weighted DiD (Stuart et al., 2014). The standard DiD method, a staple in health policy evaluations, allows for a comparison between implementation and control units. This method determines the impact of the implementation by comparing the change in outcomes at control units before and after the implementation with the corresponding change at implementation units. We derived the standard DiD estimates using maximum-likelihood repeated measures linear regression, incorporating an auto-regressive correlation structure. This approach is advantageous due to its simplicity and directness, offering a clear view of the intervention's impact. The standard DiD results are in Table S1.

| **Table S1** Standard DiD Estimates of the Impact of the National i-THRIVE Programme on THRIVE Fidelity | | | |
| --- | --- | --- | --- |
| **Outcome** | **Estimate** | **95% CI** | **p-value** |
| Overall Fidelity | 7.88 | -3.15–18.92 | 0.150 |
| Macro-level Fidelity | 3.25 | -0.61–7.10 | 0.093 |
| Meso-level Fidelity | 1.59 | -3.23–6.41 | 0.496 |
| Micro-level Fidelity | 3.05 | -1.43–7.53 | 0.169 |
| CI = confidence interval, DiD = difference in differences | | | |

The four-group propensity score weighted DiD, on the other hand, offers a more nuanced analysis. It accounts for potential imbalances between the implementation and control groups by weighting their characteristics. This method enhances the accuracy of the estimates by reducing biases that could arise from unobserved confounding variables or selection biases inherent in non-randomised studies. By comparing these two methods, we aim to validate the consistency of our findings and ensure that our conclusions are not solely dependent on a single analytical approach. Such a comprehensive analytical strategy strengthens the reliability of our conclusions regarding the effectiveness of the NIP in improving mental health care services for children and young people in England.

**Alternative Model Specification**

We included effect estimates from a Gaussian-distributed identity-link generalised linear model using generalised estimating equations with an auto-regressive correlation structure, as well as a Gamma-distributed identity-link generalised linear model using generalised estimating equations with an auto-regressive correlation structure. These results are in Table S2. Although the effect estimates from the Gamma GEE model reach a level of statistical significance for meso-level fidelity, the model fit metric (QICu) does not support selection of this model over the Gaussian GEE.

| **Table S2** Estimates of the Impact of the National i-THRIVE Programme on THRIVE Fidelity | | | | | |
| --- | --- | --- | --- | --- | --- |
| **Outcome** | **Estimator** | **QICu** | **Estimate** | **95% CI** | **p-value** |
| Overall Fidelity | Gaussian GEE | 47.00 | 6.07 | -4.43–16.56 | 0.257 |
|  | Gamma GEE | 2421.88 | 8.92 | -0.05–17.89 | 0.051 |
| Macro-level Fidelity | Gaussian GEE | 47.00 | 2.56 | -1.19–6.30 | 0.180 |
|  | Gamma GEE | 2956.39 | 3.33 | -0.24–6.90 | 0.067 |
| Meso-level Fidelity | Gaussian GEE | 47.00 | 2.85 | -0.77–6.48 | 0.123 |
|  | Gamma GEE | 1622.20 | 5.04 | 2.29–7.78 | <0.001 |
| Micro-level Fidelity | Gaussian GEE | 47.00 | 0.66 | -5.13–6.45 | 0.823 |
|  | Gamma GEE | 1712.07 | 2.22 | -3.18–7.63 | 0.420 |
| CI = confidence interval, GEE = generalized estimating equations, QICu = quasi-Akaike information criterion (u) | | | | | |

**Exclusion of Non-Compliant Control Sites**

To assess the influence of non-compliant comparison sites, we repeated the analysis while excluding Site J, the comparison site with the strongest evidence for non-compliance. These results are provided in Table S3.

| **Table S3** Estimates of the Impact of the National i-THRIVE Programme on THRIVE Fidelity while Excluding Non-compliant Comparison Sites | | | |
| --- | --- | --- | --- |
| **Outcome** | **Estimate** | **95% CI** | **p-value** |
| Overall Fidelity | 5.31 | -6.46–17.07 | 0.352 |
| Macro-level Fidelity | 2.35 | -1.77–6.47 | 0.242 |
| Meso-level Fidelity | 2.18 | -2.74–7.09 | 0.360 |
| Micro-level Fidelity | 0.79 | -4.76–6.33 | 0.767 |
| CI = confidence interval | | | |
