## Supplemental Material S8 for "Effect of a needs-based model of care on the characteristics of healthcare services in England: the i-THRIVE National Implementation Programme"

**Supplemental Material S8: Additional Results**

**Surveys**

The staff survey was distributed to 2,415 staff. Responses included 261 staff (37.9%) were from i-THRIVE sites, while 428 respondents (62.1%) were from comparison sites. The mean response rate was 33.0% at i-THRIVE sites (95% CI: 16.3–49.7%) and 34.3% at comparison sites (95% CI: 20.6–48.0%). There was no significant difference in the response rate (p = 0.9). Demographic and professional characteristics of the respondents are summarised in Table S1.

| Table S1: Staff Survey Respondents | | | |
| --- | --- | --- | --- |
|  |  | i-THRIVE | Comparison |
|  | n  (%) | 261  (37.9) | 428  (62.1) |
| Organisation | NHS Site | 220  (84.6) | 409  (96.0) |
|  | Local authority | 15  (5.8) | 7  (1.6) |
|  | Third sector | 11  (4.2) | 5  (1.2) |
|  | Education | 7  (2.7) | 1  (0.2) |
|  | Missing | 1 | 2 |
| Position | Frontline practitioner | 181  (69.6) | 297  (69.9) |
|  | Service/team manager | 47  (18.1) | 50  (11.8) |
|  | Senior lead | 18  (6.9) | 21  (4.9) |
|  | Commissioner | 5  (1.9) | 2  (0.5) |
|  | Other | 9  (3.5) | 55  (12.9) |
|  | Missing | 1 | 3 |
| Service Provided | Specialist mental health | 170  (65.1) | 339  (80.3) |
|  | Brief intervention | 29  (11.1) | 18  (4.3) |
|  | Prevention | 14  (5.4) | 13  (3.1) |
|  | Risk support | 22  (8.4) | 32  (7.6) |
|  | Triage, advice, assessment | 9  (3.5) | 6  (1.4) |
|  | Other | 17  (6.5) | 14  (3.3) |
|  | Missing | 0 | 6 |

Consistent with THRIVE principles, staff from implementation sites were from a broader range of services, with fewer from traditional specialist CYPMH services (65.1% vs. 80.3%); more respondents provided brief intervention (11.1% vs. 4.3%), prevention (5.4% vs. 3.1%), risk support (8.4% vs. 7.6%), or dedicated triage/advice/assessment (3.5% vs. 1.4%). i-THRIVE site service profiles differed from comparison sites, indicating organisational change in line with CYP needs (p<0.0001, Cramer’s V = 0.19). We examined knowledge, attitudes and behaviours between implementation and comparison sites separately (Table S2). Although respondents were broadly aware of the THRIVE Framework, those from implementation sites were significantly more likely to have heard of it (83.9% vs. 70.5%). More respondents from implementation sites reported that they used THRIVE principles personally in their daily practice (58.5% vs. 49.0%). When completing a short test on their knowledge of the five domains of the THRIVE Framework, those in implementing sites were more likely to get perfect answers (34.1% vs. 22.9%).

| **Table S2** Staff Survey Results | | | | |
| --- | --- | --- | --- | --- |
| **Question** | **i-THRIVE** | **Comparison** | **p-value** | **Cramer’s V** |
| Awareness of THRIVE | 83.9% | 70.5% | <0.0001 | 0.15 |
| Personal use of THRIVE in daily practice | 58.5% | 49.0% | 0.03 | 0.12 |
| Perfect knowledge of THRIVE Framework | 34.1% | 22.9% | 0.001 | 0.12 |

Responses from each site were compared to the mean response from all comparison sites, for questions about site implementation of THRIVE, personal use of THRIVE principles, and exhibiting perfect knowledge of THRIVE principles (Tables S3—S5). Most i-THRIVE site respondents had a higher odds of reporting site implementation of THRIVE (all except Sites C and R), compared to the comparison site average (Table S3). Some comparison site respondents also had an increased odds of reporting site implementation of THRIVE (Sites J and K), compared to the comparison site average. When reporting personal use of THRIVE principles, there were no notable differences between respondents at any site and the comparison site average, except a lower odds of reporting personal use of THRIVE principles from respondents at Site F (Table S4). Many i-THRIVE site respondents had a higher odds of exhibiting perfect knowledge of THRIVE principles (Sites B, D, G, L, and Q), compared to the comparison site average. Site J (a comparison site) respondents also had an increased odds of exhibiting perfect knowledge of THRIVE principles, compared to the comparison site average (Table S5).

| **Table S3** Odds of Responders at Sites Reporting Site Implementation of THRIVE | | | |
| --- | --- | --- | --- |
| **Site Type** | **Sites** | **Odds Ratio** | **95% CI** |
| i-THRIVE | Site B | – | – |
|  | Site C | 0.59 | 0.10–3.37 |
|  | Site D | 4.71 | 1.76–12.57 |
|  | Site G | 8.83 | 1.91–40.77 |
|  | Site L | 4.51 | 1.69–12.09 |
|  | Site O | 4.91 | 1.84–13.05 |
|  | Site Q | 4.20 | 1.86–10.66 |
|  | Site R | 1.18 | 0.43–3.23 |
|  | Site T | 11.77 | 1.45–95.67 |
| Comparison | Site A | 0.51 | 0.19–1.40 |
|  | Site E | 0.82 | 0.24–2.82 |
|  | Site F | 0.35 | 0.07–1.85 |
|  | Site H | 3.07 | 0.74–12.71 |
|  | Site I | 0.31 | 0.08–1.23 |
|  | Site J | 4.43 | 2.32–8.47 |
|  | Site K | 4.43 | 1.33–14.80 |
|  | Site N | 0.55 | 0.17–1.77 |
|  | Site P | 1.78 | 0.78–4.04 |
|  | Site S | 0.35 | 0.04–3.51 |
| Referent | Mean of all comparison sites | 1.00 | – |
| * all respondents from Site B responded “yes”, thus an odds ratio could not be calculated  CI = confidence interval, | | | |

| **Table S4** Odds of Responders at Sites Reporting Personal Use of THRIVE Principles | | | |
| --- | --- | --- | --- |
| **Site Type** | **Site** | **Odds Ratio** | **95% CI** |
| i-THRIVE | Site B | – | – |
|  | Site C | 0.30 | 0.06–1.43 |
|  | Site D | 0.96 | 0.35–2.61 |
|  | Site G | 1.71 | 0.45–6.56 |
|  | Site L | 1.58 | 0.53–4.70 |
|  | Site O | 1.25 | 0.44–2.52 |
|  | Site Q | 0.92 | 0.37–2.40 |
|  | Site R | 0.87 | 0.27–2.77 |
|  | Site T | 1.58 | 0.31–8.00 |
| Comparison | Site A | 1.03 | 0.32–3.29 |
|  | Site E | 1.89 | 0.38–9.54 |
|  | Site F | 0.17 | 0.03–0.97 |
|  | Site H | 2.61 | 0.30–22.90 |
|  | Site I | 0.99 | 0.28–3.52 |
|  | Site J | 1.23 | 0.60–2.51 |
|  | Site K | 1.89 | 0.38–9.54 |
|  | Site N | 1.21 | 0.30–4.93 |
|  | Site P | 0.56 | 0.20–1.58 |
|  | Site S | 0.77 | 0.07–8.88 |
| Referent | Mean of all comparison sites | 1.00 | – |
| * all respondents from Site B responded “yes”, thus an odds ratio could not be calculated  CI = confidence interval | | | |

| **Table S5** Odds of Responders at Sites Having Perfect Knowledge of THRIVE  Perfect knowledge was defined as achieving a perfect score on the THRIVE quiz. | | | |
| --- | --- | --- | --- |
| **Site Type** | **Site** | **Odds Ratio** | **95% CI** |
| i-THRIVE | Site B | 3.60 | 1.21–10.70 |
|  | Site C | 1.54 | 0.31–7.71 |
|  | Site D | 2.70 | 1.27–5.72 |
|  | Site G | 2.70 | 1.02–7.13 |
|  | Site L | 5.06 | 2.30–11.10 |
|  | Site O | 1.90 | 0.90–4.01 |
|  | Site Q | 11.24 | 5.19–24.31 |
|  | Site R | 0.75 | 0.30–1.89 |
|  | Site T | 1.47 | 0.39–5.53 |
| Comparison | Site A | 0.45 | 0.15–1.35 |
|  | Site E | 0.34 | 0.10–1.15 |
|  | Site F | 0.69 | 0.15–3.16 |
|  | Site H | 0.65 | 0.14–2.92 |
|  | Site I | 1.39 | 0.59–3.30 |
|  | Site J | 3.73 | 2.25–6.19 |
|  | Site K | 1.15 | 0.37–3.60 |
|  | Site N | 1.92 | 0.71–5.20 |
|  | Site P | 1.57 | 0.71–3.48 |
|  | Site S | 0.81 | 0.18–3.75 |
| Referent | Mean of all comparison sites | 1.00 | – |
| CI = confidence interval | | | |

**THRIVE Fidelity**

For fidelity scores, the interrater reliability (Krippendorf’s alpha) among the raters ranged from 0.57 to 0.93, with a mean alpha of 0.73.

Unweighted group characteristics are in Table S6, with the weighted standardised difference in means. An important indicator of balanced group characteristics is a standardised difference in means of less than 0.25 for all characteristics and groups, with the pre-implementation i-THRIVE group serving as the baseline for comparison.

| **Table S6** Four-Group Propensity Score Weighted Characteristics of i-THRIVE and Comparison Sites | | | | | | | | |
| --- | --- | --- | --- | --- | --- | --- | --- | --- |
| **Characteristics** | **Mean (SD)** | | | | | **Weighted standardised difference in means, compared to Pre i-THRIVE** | | |
|  | **i-THRIVE** | | **Control** | | | **Post i-THRIVE** | **Pre control** | **Post control** |
|  | **Pre**  (n=10) | **Post**  (n=10) | | **Pre**  (n=10) | **Post**  (n=10) |  |  |  |
| Population density (persons per 10 km^2^) | 49.7  (47.2) | 52.2  (51.5) | | 23.9  (29.0) | 24.5  (29.8) | 0.061 | 0.253 | 0.165 |
| Funding  (£100,000) | 47.3  (47.4) | 60.4  (41.4) | | 44.3  (21.1) | 56.2  (34.9) | 0.018 | 0.210 | 0.067 |
| IMD | 84.0  (60.4) | 93.8  (51.8) | | 69.1  (44.7) | 67.8  (45.5) | 0.056 | 0.292 | 0.256 |
| Baseline Number of CCGs | 1.4  (1.0) | 1.4  (1.0) | | 1.7  (1.3) | 1.7  (1.3) | 0.039 | 0.000 | 0.200 |
| Transformation progress | 72.0  (27.0) | 72.0  (27.0) | | 64.0  (20.0) | 64.0  (20.0) | 0.108 | 0.265 | 0.383 |
| CCG = Clinical Commissioning Group, IMD = Indices of Multiple Deprivation, Post = post-implementation, Pre = pre-implementation, SD = standard deviation | | | | | | | | |
