## Supplemental Material S9 for "Effect of a needs-based model of care on the characteristics of healthcare services in England: the i-THRIVE National Implementation Programme"

**Supplemental Material S9: i-THRIVE Effect Moderation**

This section of the supplemental materials details the approach used to investigate the influence of effective working relationships among local systems on the impact of the i-THRIVE programme in enhancing fidelity.

**Working Relationships as a Moderating Factor**

The i-THRIVE model emphasizes the importance of strong working relationships at various levels. At the macro level, it encourages cooperation among various agencies, such as educational and social services, in policy making and care commissioning (Moore et al., 2023). At the meso level, a THRIVE-aligned site would typically feature a network of community providers (Moore et al., 2023). Achieving these objectives is significantly influenced by the effectiveness of working relationships at the site as well as within the broader community, including supporting agencies. These collaborations are essential for the successful implementation of i-THRIVE and are largely contingent upon the willingness and interest of these agencies to participate in CAMHS activities.

Given this context, we explored the role of effectiveness of working relationships among local systems. Our hypothesis was that effectiveness of working relationships would likely affect macro- and meso-level fidelity scores and potentially the overall fidelity score, though not on micro-level fidelity scores.

**Data and Methods**

We utilized the four-group propensity-score weights derived from the sensitivity analyses (see Supplementary Materials S7). To gauge effectiveness of local system working relationships at each site, we used the "effectiveness of local system working relationships" component rating from the CCG Assurance Annual Assessment 2017/18 (NHS England, 2021). We averaged these ratings for sites with multiple CCGs. A threshold of 68.0 was set to distinguish sites with highly effective local system working relationships. The mean overall score was 68.9 for comparison sites and 68.3 for implementation sites, with five highly effective comparison sites (Sites E, I, N, P, and S) and five highly effective implementation sites (Sites D, L, M, O, and T) identified.

We employed a difference-in-difference-in-differences (DiDiD) model to assess i-THRIVE's impact on macro-level fidelity. This analysis was conducted using maximum-likelihood repeated measures linear regression with an auto-regressive correlation structure, weighted with propensity scores. The model included a three-way interaction of variables representing highly effective working relationships, the post-implementation period, and the intervention group. To account for any remaining differences in characteristics, we included the covariates population density, IMD rank, and transformation compliance in the final model. This methodological approach allows us to discern the moderating effect of working relationship effectiveness on the impact of i-THRIVE, providing a more comprehensive understanding of the factors contributing to successful implementation.
